# LIVER-RELATED AND OTHER-CAUSE MORTALITY RISK IN EXCESSIVE DRINKERS: ANALYSIS OF DATA FROM THE UK BIOBANK

**DOI:** 10.1101/2022.06.01.22275852

**Authors:** John B Whitfield, Devanshi Seth, Timothy R Morgan, GenomALC Consortium

## Abstract

**Background and Aims:** High alcohol intake is associated with increased mortality. We aimed to identify factors affecting mortality in people drinking extreme amounts.

**Approach and Results:** Information was obtained from the UK Biobank on approximately 500,000 participants aged 40-70 years at baseline assessment in 2006-2010. Habitual alcohol intake, lifestyle and physiological data, laboratory test results, and hospital diagnoses and death certificate data (to June 2020) for 5136 men (2.20% of male participants) and 1504 women (0.60%) who reported taking ≥80 g/day or ≥50g/day, respectively, were used in survival analysis. Compared to all other participants, their mortality HRs were 9.40 (95% CI 7.00-12.64) for any liver disease (ICD-10 K70-K76), 2.02 (1.89-2.17) for all causes, 1.89 (1.69-2.12) for any cancer (C00-C99), and 1.87 (1.61-2.17) for any circulatory disease (I00-I99). Liver disease diagnosis or abnormal liver function tests predicted not only deaths attributed to liver disease but also those from cancers or circulatory diseases. Mortality among excessive drinkers was also associated with quantitative alcohol intake, diagnosed alcohol dependence (ICD-10 F10.2), and current smoking at assessment.

**Conclusions:** People with chronic excessive alcohol intake experience decreased average survival but there is substantial variation in their mortality, with liver abnormality and alcohol dependence each associated with worse prognosis. Clinically, patients with these risk factors as well as high alcohol intake should be considered for early or intensive management. Research can usefully focus on the factors predisposing to dependence or liver abnormality.

## INTRODUCTION

Many studies have compared all-cause mortality, mortality from pre-defined causes, or incidence of conditions of interest between groups of people classified by their self-reported alcohol intake (1-4). Most have found associations between higher alcohol intake and poorer outcomes. Few studies have asked the complementary question -what happens to people who do consume hazardous amounts of alcohol? How much are their lives shortened, what are their causes of death, and what factors modify or predict mortality and morbidity in people with persistently high alcohol intake? There have been estimates of alcohol-related disease burden based on prevalence of high intake and the dose-response curve for mortality (5), and studies based on follow-up of patients with alcohol dependence (6-10), but information on long-term outcomes for excessive drinkers from the general population is scarce. A direct approach to these questions requires prospective study of a substantial cohort of high-risk drinkers, representative of the general population and not biased towards patients seeking treatment.

Such studies require an operational definition of ‘excessive’ drinking. Reported consumption of 80 grams of alcohol per day for men, 50 grams per day or more for women, is frequently associated with adverse consequences (1, 11, 12) (although lower limits are appropriate for public health messaging and for reduction of alcohol-related risk across the population). The UK Biobank is a prime source of relevant information for the general population of an economically developed country. This project recruited just over 500,000 UK-resident men and women, aged 40-70 years in 2006-2010, and the resulting resource (13) contains self-report information on alcohol use and on a wide range of personal, physiological, biochemical and genetic characteristics. Subsequent information from death certificates and hospital records allows comparison of outcomes for excessive drinkers versus the rest of the participating population. Because the study sample is so large, it contains a substantial number of excessive drinkers even though they comprise only a small proportion of participants.

Our analysis of the UK Biobank data addresses questions related to risk in excessive drinkers. How do their all-cause mortality, and the distribution of causes, compare with those who do not meet excessive drinking criteria? In particular, how prominent is alcohol-related liver disease (recognised as a major medical consequence of excessive drinking), or liver disease in general, among the causes of death? What risk factors or predictors are associated with risk of death among excessive drinkers, and how large are their effects?

## SUBJECTS AND METHODS

Information used in this analysis was obtained from the UK Biobank (application number 18870). Recruitment and initial assessment of just over 500,000 participants occurred between 2006 and 2010, targeted at the 40-69 years age group (actual range 37-73, median 58 years). Participants gave informed consent, and our application was approved by the UK Biobank through their procedures, consistent with the UK Biobank Ethics and Governance Framework (https://www.ukbiobank.ac.uk/learn-more-about-uk-biobank/about-us/ethics, accessed 2022-03-22). A small number of participants who had withdrawn their consent after initial participation were excluded from the data analysis.

Information obtained at an assessment session included responses to computer-administered questionnaires, physiological measurements and collection of blood samples for laboratory tests and genotyping. Questions about alcohol use included quantity and frequency of specified alcoholic beverages at the time of assessment, and a simple comparison with the amount ten years previously. Participants’ answers were used to calculate the number of standard drinks per week and hence the amount of alcohol in grams per day. Initial assessment included information about smoking, body mass index (BMI), blood pressure and consumption of non-alcoholic beverages including tea and coffee. Details are available at https://biobank.ndph.ox.ac.uk/showcase/ (accessed 2022-03-22).

Excessive drinkers were defined, for this analysis, as participants with self-reported alcohol intake at the time of assessment greater than or equal to 80 g/day for men, 50 g/day for women and also (to ensure that they had long-term high exposure) similar or greater consumption ten years previously.

Information on diagnoses from UK hospital encounters, and from death certificates, was coded using the International Classification of Diseases, 10^th^ Edition (ICD-10). Death records were obtained in June 2020 and dates of death ranged from 10 May 2006 to 26 April 2020.

IBM SPSS Statistics, version 22, was used for data management and statistical tests. The main results are based on survival analysis using either Kaplan-Meier or Cox regression methods. Because participants entered the study at differing ages and times, and exposure to alcohol-related risk was over a substantial time, age at death was used as the survival variable. Except where comparisons were made between results for men and women, sex was included as a factor in survival analyses. Analysis based on all-cause mortality was supplemented with more detailed analysis by cause of death, using only the underlying (primary) cause of death (UK Biobank field 40001). Sub-groups were created for deaths from all cancers (ICD-10 C00 to C99), all circulatory diseases (ICD-10 I00 to I99), and all other causes except liver disease. Because of its strong association with alcohol use, there was a separate analysis for deaths from any liver disease (ICD-10 K70 to K76, including both alcohol-related liver disease and other liver diseases). Age at death was calculated from the year of birth (exact dates of birth are not available) and the date of death, and participants who had not died were censored at an age calculated from the difference between year of birth and 2020.

Frequency distributions for laboratory test results were plotted and examined visually; if appropriate the results were log_10_-transformed to obtain more symmetrical distributions. Data (for excessive drinkers only) were regressed on age at blood collection, separately for men and women, and the standardised residuals were saved for use as predictors in the survival analysis. Amount of alcohol reported by the excessive drinkers was also log-transformed. Binary variables such as presence or absence of a diagnosis were coded as 0 (absent) or 1 (present).

## RESULTS

### Comparisons between excessive-drinking and other participants

Among the UK Biobank participants, 5136 men (2.20% of male participants) reported drinking at or above 80 g of alcohol/day with similar or greater consumption 10 years previously, and 1504 women (0.60% of female participants) reported drinking 50 g/day or more at the time of assessment with similar or greater consumption 10 years earlier. These participants are referred to as the ‘excessive drinkers’. Descriptive information for excessive drinkers, and for all other participants, is shown in Supplementary Table 1.

811 (12.2%) of the 6640 excessive drinkers and 29,448 (5.9%) of the other participants were recorded as having died from any cause by June 2020. Survival analysis showed a sex-adjusted Hazard Ratio of 2.02 (95% CI 1.89-2.17, p = 2.54 × 10^−86^) for all-cause mortality among excessive drinkers, similar for men (1.99, 1.85-2.15) and women (2.09, 1.73-2.52). Estimated mean survival was 79.2 years for excessive drinkers and 83.1 years for all others in men, 80.4 and 82.5 years respectively in women (Figure 1). Results of survival analysis using only those who reported alcohol intake of 5-15 g/day as the comparison group, rather than all participants who did not meet the criteria for excessive drinking, gave slightly higher HRs (Supplementary Table 2).

**Figure 1.**
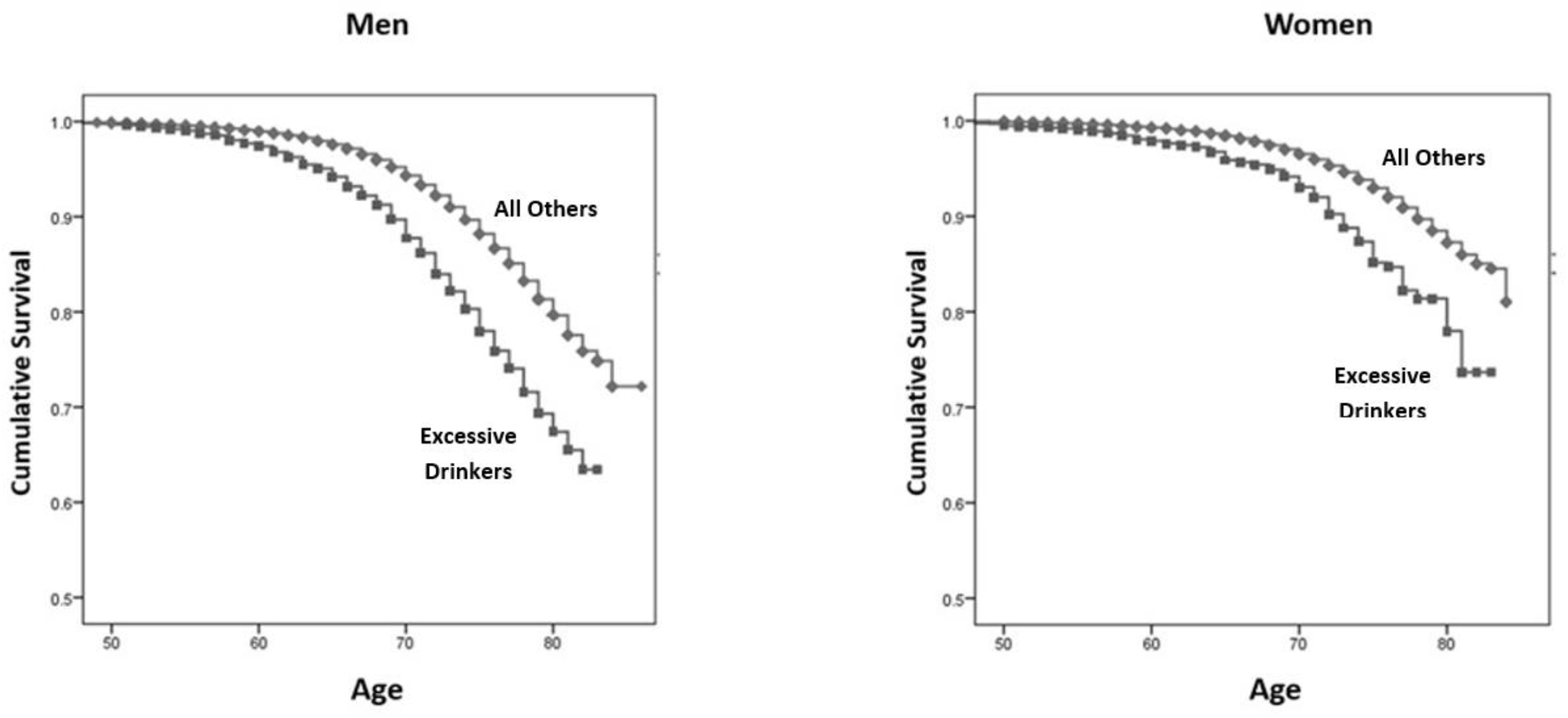
Kaplan-Meier survival analysis showing effects of excessive drinking on all-cause mortality. The lines contrast survival for the excessive drinkers and for all others. For men the difference in mean estimated age at death was 3.9 years (79.2 versus 83.1, p = 8.42 × 10^−75^) and for women 2.1 years (80.4 versus 82.5, p = 2.94 × 10^−15^).

The proportions of total deaths and estimated Hazard Ratios (HR) by major ICD-10 cause-of death groups, comparing excessive drinkers against all other participants, are shown in Figure 2 and Supplementary Table 3. Information on how these HR estimates change when other risk factors were added, firstly smoking status and then body mass index (BMI), waist/hip ratio (WHR), years of education, self-reported diabetes at the time of assessment, and systolic and diastolic blood pressures (SBP, DBP) are shown in Supplementary Table 4.

**Figure 2.**
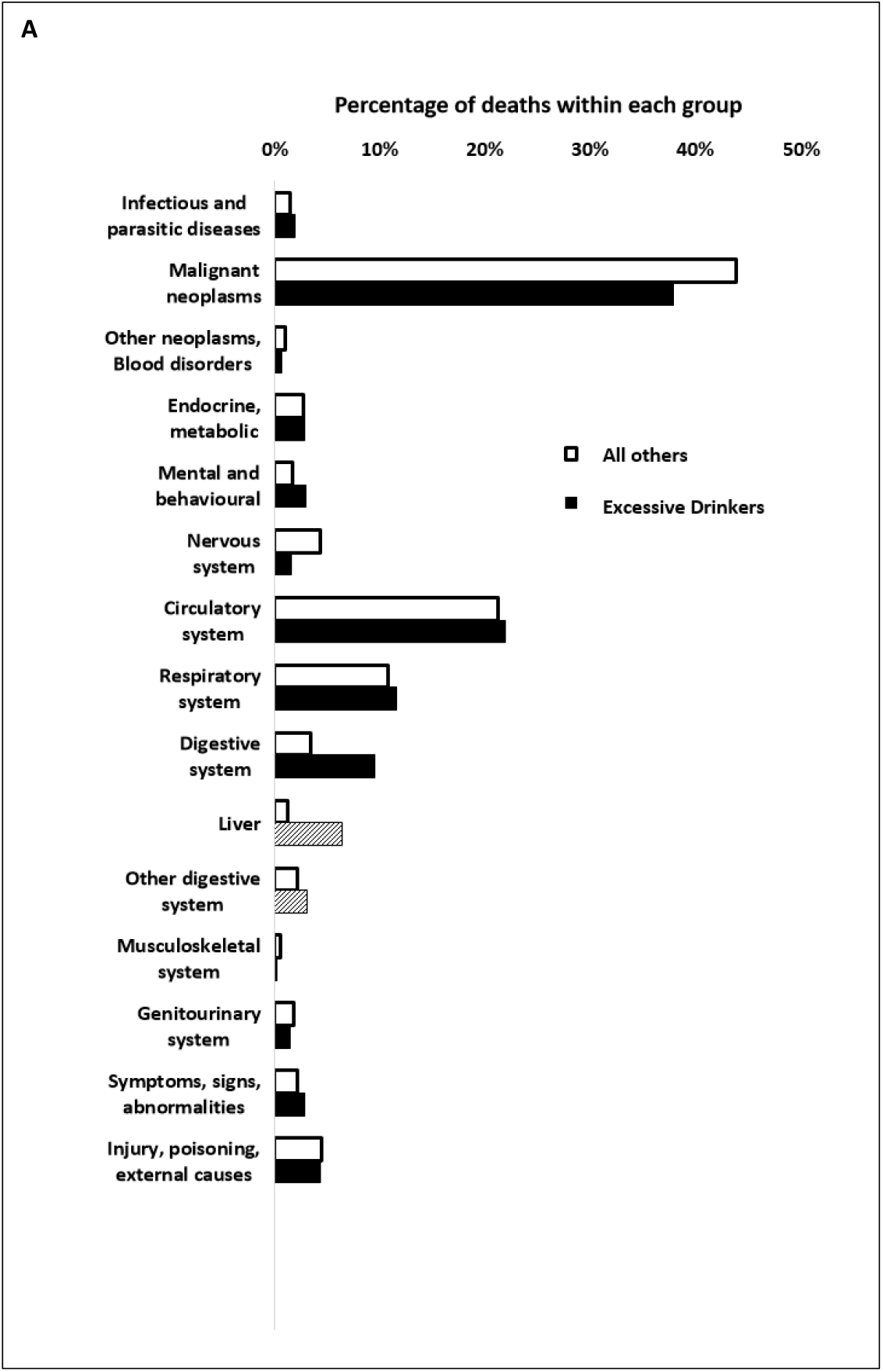
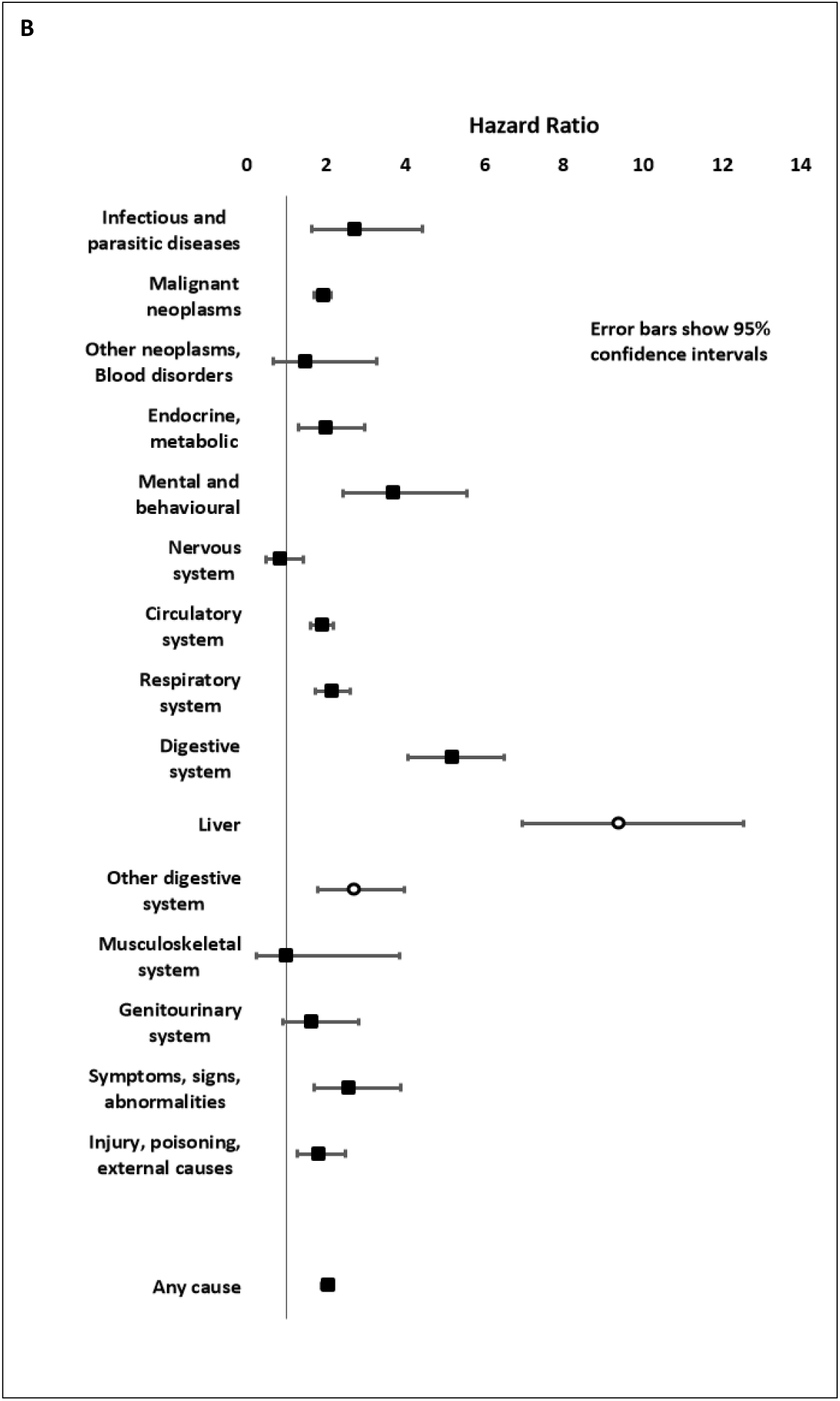
Comparison of causes of death by major ICD-10 category in excessive drinkers and all other UK Biobank participants (panel A), and Hazard Ratios for excessive drinkers compared to others by cause-of-death category (panel B). The vertical line in panel B indicates a Hazard ratio of 1.0. Results for deaths attributed to digestive system diseases (ICD-10 K00 to K93) are further divided into those from liver disease (K70 to K76) and from other disgestive disease (shown by lighter shading and error bars).

Deaths attributed to diseases of the digestive system were more common among the excessive drinkers (9.5% of deaths) compared to other participants (3.4%), with the excess mostly due to liver diseases. Other causes of death were similar between the two groups (Figure 2, panel A). The highest proportion of deaths in both groups were from cancers and circulatory diseases. The HRs for most cause-of-death groups were in the range 1.5 to 2.5, consistent with our all-cause mortality estimate of 2.0. However liver disease (HR 9.34, 95% CI 6.95-12.56) and mental and behavioural disorders (3.68, 2.44-5.56) had higher, and nervous system diseases lower (0.82 [0.48-1.42]), HRs (Figure 2, panel B; Supplementary Table 3).

### Liver disease in excessive drinkers

Because of the important association between alcohol and liver disease we firstly considered deaths ascribed to any liver disease (including but not limited to alcohol-related liver disease). Fifty-two out of 6640 (0.78%) excessive drinkers and 355 out of 496,036 (0.07%) other participants died of liver diseases between the time of recruitment and June 2020.

Variation in liver disease mortality among excessive drinkers was associated with multiple risk factors (Tables 1 and 2). Smoking and alcohol-related conditions including current smoking (at the time of assessment), amount of alcohol, and diagnosis of alcohol dependence were each associated with liver deaths but when these factors were considered together (Table 1, lower part) alcohol dependence was the most significant.

**Table 1.**
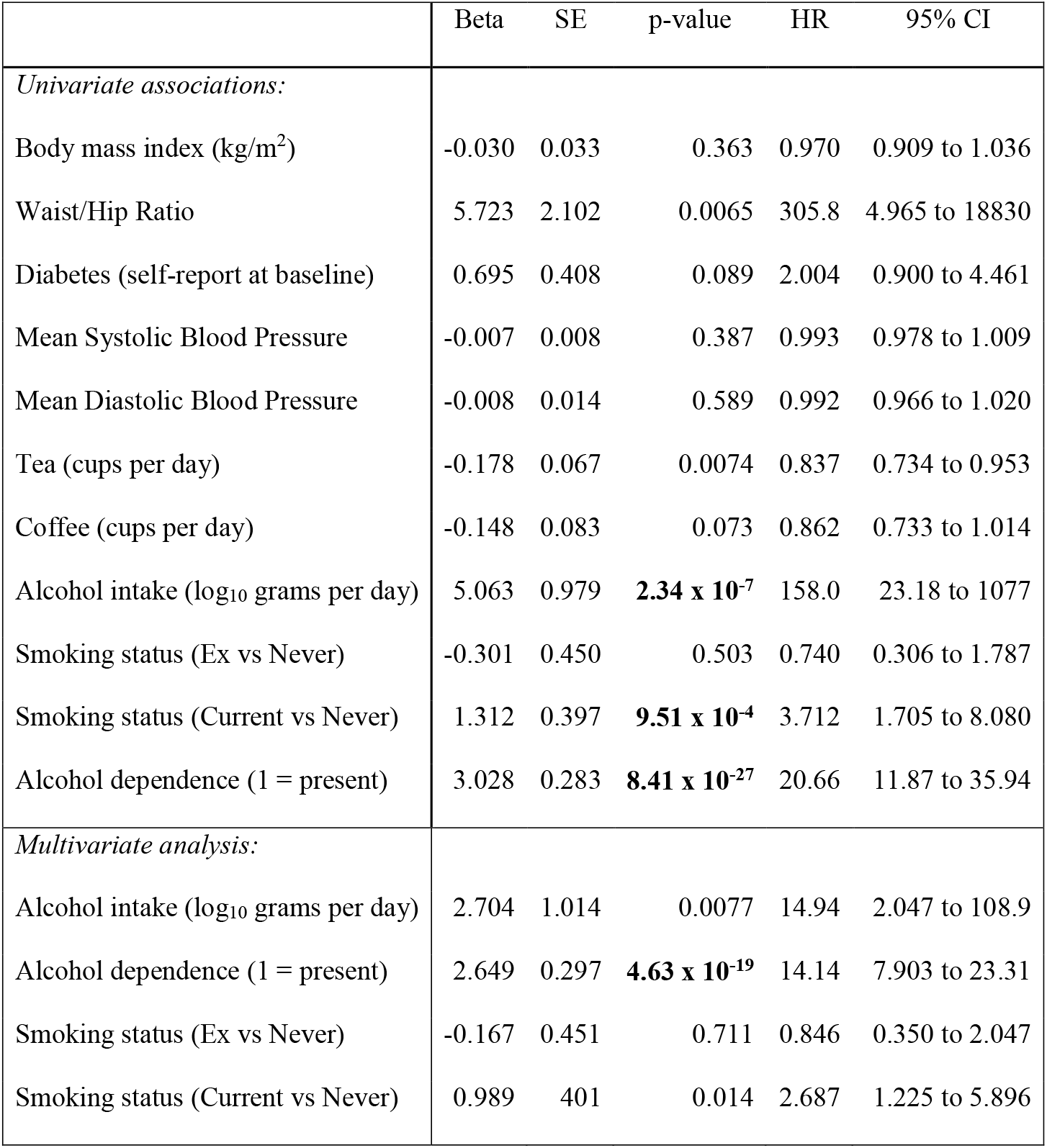
Risk Factors affecting sex-adjusted mortality from any liver disease (ICD-10 codes K70 to K77) in excessive drinkers. Results in the upper section (univariate associations) are not adjusted for the effects of the other factors listed, while the results in the lower section (multivariate analysis) estimate the independent effects of intake, dependence and smoking (adjusted for each other).

|  | Beta | SE | p-value | HR | 95% CI |
| --- | --- | --- | --- | --- | --- |
| *Univariate associations:* |  |  |  |  |  |
| Body mass index (kg/m<sup>2</sup>) | -0.030 | 0.033 | 0.363 | 0.970 | 0.909 to 1.036 |
| Waist/Hip Ratio | 5.723 | 2.102 | 0.0065 | 305.8 | 4.965 to 18830 |
| Diabetes (self-report at baseline) | 0.695 | 0.408 | 0.089 | 2.004 | 0.900 to 4.461 |
| Mean Systolic Blood Pressure | -0.007 | 0.008 | 0.387 | 0.993 | 0.978 to 1.009 |
| Mean Diastolic Blood Pressure | -0.008 | 0.014 | 0.589 | 0.992 | 0.966 to 1.020 |
| Tea (cups per day) | -0.178 | 0.067 | 0.0074 | 0.837 | 0.734 to 0.953 |
| Coffee (cups per day) | -0.148 | 0.083 | 0.073 | 0.862 | 0.733 to 1.014 |
| Alcohol intake (log<sub>10</sub> grams per day) | 5.063 | 0.979 | **2.34 x 10<sup>-7</sup>** | 158.0 | 23.18 to 1077 |
| Smoking status (Ex vs Never) | -0.301 | 0.450 | 0.503 | 0.740 | 0.306 to 1.787 |
| Smoking status (Current vs Never) | 1.312 | 0.397 | **9.51 x 10<sup>-4</sup>** | 3.712 | 1.705 to 8.080 |
| Alcohol dependence (1 = present) | 3.028 | 0.283 | **8.41 x 10<sup>-27</sup>** | 20.66 | 11.87 to 35.94 |
| *Multivariate analysis:* |  |  |  |  |  |
| Alcohol intake (log<sub>10</sub> grams per day) | 2.704 | 1.014 | 0.0077 | 14.94 | 2.047 to 108.9 |
| Alcohol dependence (1 = present) | 2.649 | 0.297 | **4.63 x 10<sup>-19</sup>** | 14.14 | 7.903 to 23.31 |
| Smoking status (Ex vs Never) | -0.167 | 0.451 | 0.711 | 0.846 | 0.350 to 2.047 |
| Smoking status (Current vs Never) | 0.989 | 401 | 0.014 | 2.687 | 1.225 to 5.896 |

**Table 2.**
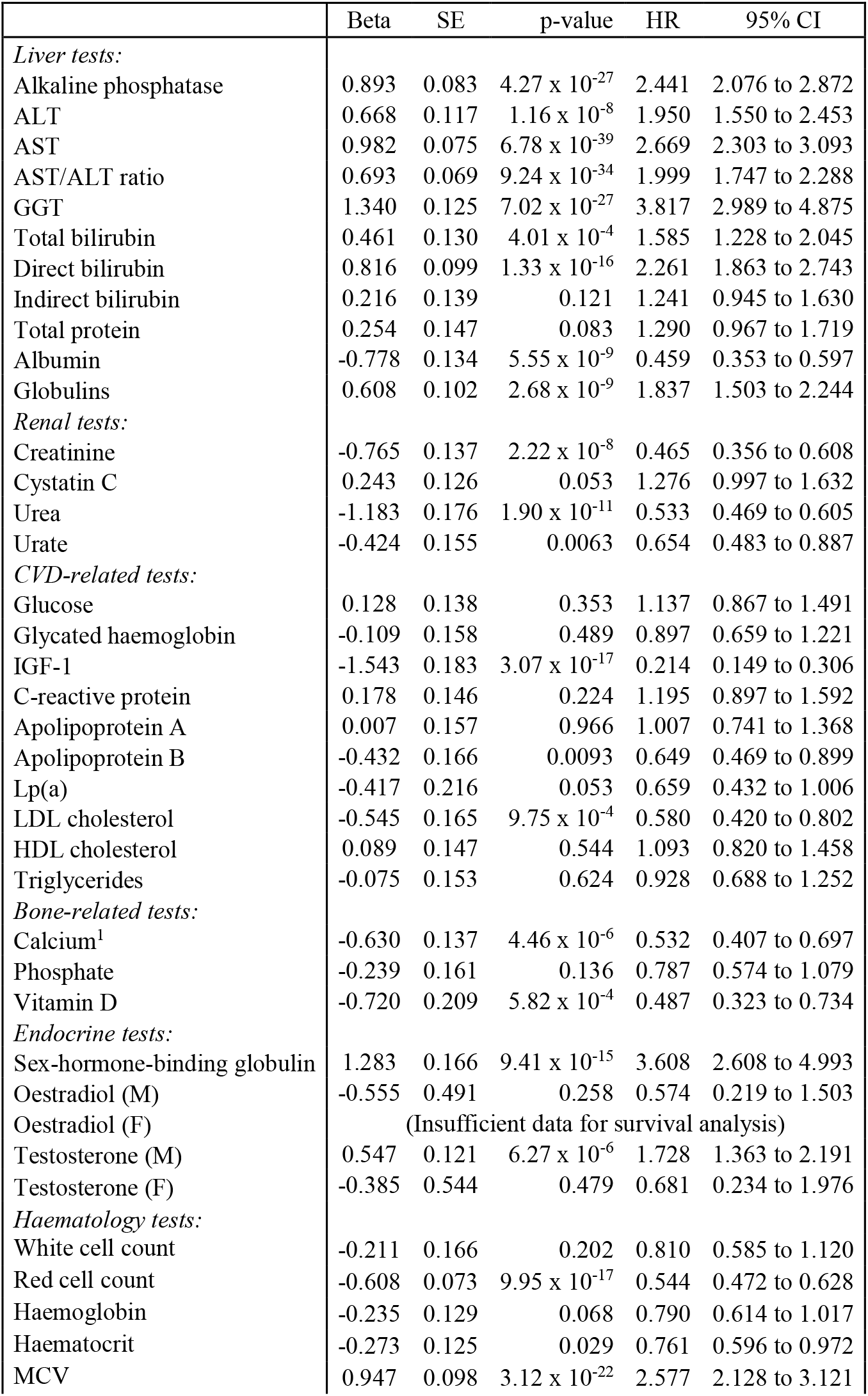
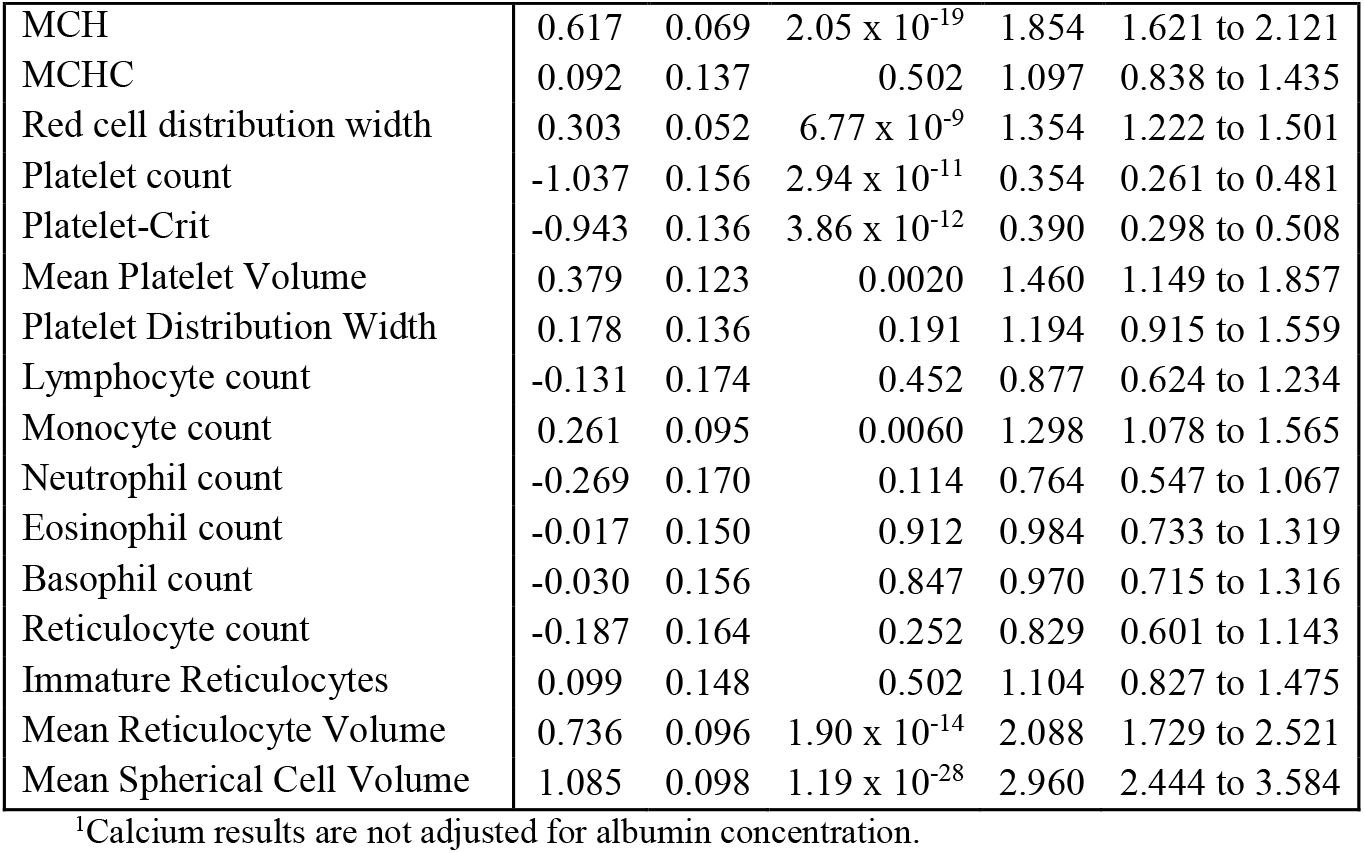
Mortality for deaths ascribed to any liver disease (ICD-10 codes K70 to K77) in excessive drinkers, by age- and sex-adjusted standardised laboratory test results.

Many of the laboratory test results from the baseline assessment showed significant associations with risk of death from liver disease (Table 2). The liver function tests gamma-glutamyl transferase (GGT), aspartate aminotransferase (AST), alkaline phosphatase and direct bilirubin each showed doubling or more of risk for each 1-Standard Deviation increase in the age- and sex-adjusted standardised residuals for the excessive drinkers group. For renal tests, creatinine, urea and uric acid were inversely associated with risk but cystatin C was not. There were also strong positive associations (higher values = higher risk) between deaths from liver disease and sex hormone binding globulin (SHBG) and testosterone (in men only); and negative associations (higher values = lower risk) for insulin-like growth factor-1 (IGF-1) and vitamin D. Associations with haematology tests were mostly related to erythrocyte size or shape, or to platelet count.

### Overall mortality in the excessive-drinking group

Few of the risk factors expected to affect all-cause mortality in the general population were statistically significant in the excessive-drinkers group (Table 3).Diagnosed alcohol-related liver disease (ICD-10 K70.0 to K70.9) was associated with increased all-cause mortality.

**Table 3.** Risk Factors affecting sex-adjusted all-cause mortality in excessive drinkers. Results in the upper section (univariate associations) are not adjusted for the effects of the other factors listed, while the results in the lower section (multivariate analysis) estimate the independent effects of intake, dependence and smoking (adjusted for each other).

|  | Beta | SE | p-value | HR | 95% CI |
| --- | --- | --- | --- | --- | --- |
| *Univariate associations:* |  |  |  |  |  |
| Body mass index (kg/m<sup>2</sup>) | -0.031 | 0.009 | 2.44 x 10<sup>-4</sup> | 0.969 | 0.953 to 0.986 |
| Waist/Hip Ratio | 1.643 | 0.557 | 0.0032 | 5.169 | 1.735 to 15.399 |
| Diabetes (self-report at baseline) | 0.242 | 0.121 | 0.046 | 1.273 | 1.004 to 1.614 |
| Mean Diastolic Blood Pressure | -0.007 | 0.004 | 0.060 | 0.993 | 0.986 to 1.000 |
| Mean Systolic Blood Pressure | -0.005 | 0.002 | 0.013 | 0.995 | 0.991 to 0.999 |
| Tea (cups per day) | -0.015 | 0.013 | 0.232 | 0.985 | 0.961 to 1.010 |
| Coffee (cups per day) | 0.011 | 0.017 | 0.539 | 1.011 | 0.977 to 1.045 |
| Alcohol intake (log<sub>10</sub> grams per day) | 2.373 | 0.319 | 9.72 x 10<sup>-14</sup> | 10.73 | 5.74 to 20.03 |
| Smoking status (Ex vs Never) | 0.147 | 0.106 | 0.166 | 1.158 | 0.941 to 1.425 |
| Smoking status (Current vs Never) | 1.062 | 0.105 | 5.47 x 10<sup>-24</sup> | 2.892 | 2.353 to 3.553 |
| Alcohol dependence (1 = present) | 1.565 | 0.087 | 9.33 x 10<sup>-72</sup> | 4.784 | 4.031 to 5.678 |
| Any alcoholic liver disease, K70.0-K70.9 (1 = present) | 1.969 | 0.099 | 9.41 x 10<sup>-89</sup> | 7.166 | 5.907 to 8.694 |
| *Multivariate analysis:* |  |  |  |  |  |
|  | Beta | SE | p-value | HR | 95% CI |
| Alcohol intake (log<sub>10</sub> grams per day) | 1.452 | 0.324 | 7.25 x 10<sup>-6</sup> | 4.274 | 2.266 to 8.061 |
| Alcohol dependence (1 = present) | 1.337 | 0.091 | 2.77 x 10<sup>-49</sup> | 3.807 | 3.188 to 4.547 |
| Smoking status (Ex vs Never) | 0.179 | 0.106 | 0.091 | 1.196 | 0.972 to 1.472 |
| Smoking status (Current vs Never) | 0.951 | 0.106 | 2.29 x 10<sup>-19</sup> | 2.588 | 2.104 to 3.183 |

Other factors related to alcohol use including reported amount of alcohol (even though results in this Table are for people meeting the excessive drinking criteria); smoking; alcohol dependence (ICD-10 F10.2, mental and behavioural disorders due to use of alcohol; dependence syndrome) each had highly significant detrimental associations with survival. For smoking, the major difference was between current and never/former smokers (at the time of assessment). Because these substance-use measures are strongly associated an analysis including alcohol consumption, alcohol dependence and smoking was conducted, with results shown in the second part of Table 3. Each remained significant but diagnosis of alcohol dependence showed the most significant association. The effect size for amount of alcohol appears large in relation to the other factors included but, because this measure was log-transformed, the HR of 4.27 in the multivariate analysis applies to a ten-fold increase in alcohol intake. An alternative analysis, comparing excessive drinkers with alcohol intake above versus below the sex-specific medians for this group, showed HR = 1.17 (1.02-1.35) in a multivariate analysis with alcohol dependence and smoking status.

More detailed analysis of risk factors, with separation of causes of death into cancers, circulatory diseases and other causes, is shown in Table 4. Cancer deaths in excessive drinkers were affected by smoking and alcohol dependence but not amount of alcohol. Circulatory system deaths were also affected by these factors, and by diagnosed alcohol-related liver disease. Mortality from the remaining causes of death except liver disease was associated with smoking, amount of alcohol consumed, alcohol dependence and reported alcohol-related liver disease (Table 4).

**Table 4.**
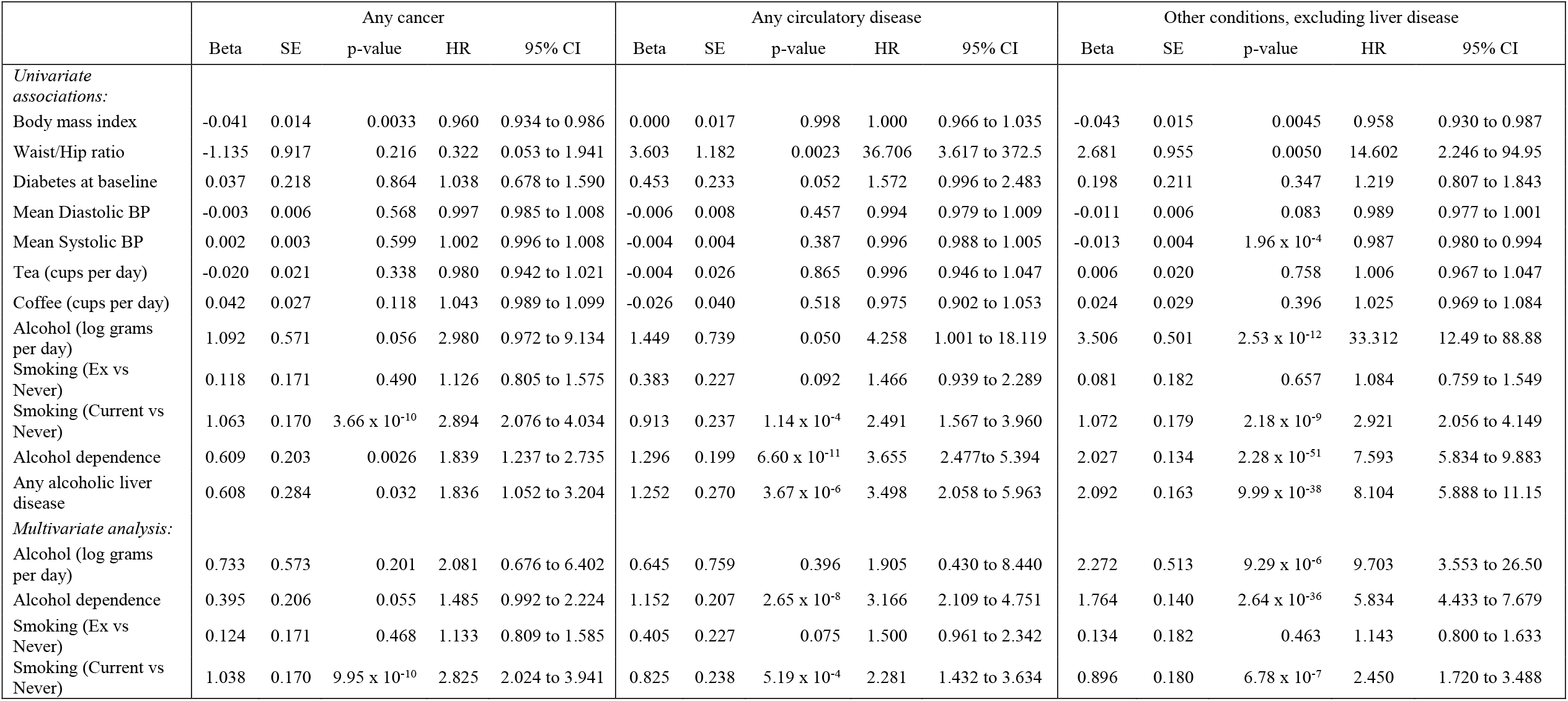
Risk Factors affecting sex-adjusted mortality, by major cause-of-death groups, in

|  | Any cancer |  |  |  |  | Any circulatory disease |  |  |  |  | Other conditions, excluding liver disease |  |  |  |  |
| --- | --- | --- | --- | --- | --- | --- | --- | --- | --- | --- | --- | --- | --- | --- | --- |
|  | Beta | SE | p-value | HR | 95% CI | Beta | SE | p-value | HR | 95% CI | Beta | SE | p-value | HR | 95% CI |
| *Univariate associations:* |  |  |  |  |  |  |  |  |  |  |  |  |  |  |  |
| Body mass index | -0.041 | 0.014 | 0.0033 | 0.960 | 0.934 to 0.986 | 0.000 | 0.017 | 0.998 | 1.000 | 0.966 to 1.035 | -0.043 | 0.015 | 0.0045 | 0.958 | 0.930 to 0.987 |
| Waist/Hip ratio | -1.135 | 0.917 | 0.216 | 0.322 | 0.053 to 1.941 | 3.603 | 1.182 | 0.0023 | 36.706 | 3.617 to 372.5 | 2.681 | 0.955 | 0.0050 | 14.602 | 2.246 to 94.95 |
| Diabetes at baseline | 0.037 | 0.218 | 0.864 | 1.038 | 0.678 to 1.590 | 0.453 | 0.233 | 0.052 | 1.572 | 0.996 to 2.483 | 0.198 | 0.211 | 0.347 | 1.219 | 0.807 to 1.843 |
| Mean Diastolic BP | -0.003 | 0.006 | 0.568 | 0.997 | 0.985 to 1.008 | -0.006 | 0.008 | 0.457 | 0.994 | 0.979 to 1.009 | -0.011 | 0.006 | 0.083 | 0.989 | 0.977 to 1.001 |
| Mean Systolic BP | 0.002 | 0.003 | 0.599 | 1.002 | 0.996 to 1.008 | -0.004 | 0.004 | 0.387 | 0.996 | 0.988 to 1.005 | -0.013 | 0.004 | 1.96 x 10<sup>-4</sup> | 0.987 | 0.980 to 0.994 |
| Tea (cups per day) | -0.020 | 0.021 | 0.338 | 0.980 | 0.942 to 1.021 | -0.004 | 0.026 | 0.865 | 0.996 | 0.946 to 1.047 | 0.006 | 0.020 | 0.758 | 1.006 | 0.967 to 1.047 |
| Coffee (cups per day) | 0.042 | 0.027 | 0.118 | 1.043 | 0.989 to 1.099 | -0.026 | 0.040 | 0.518 | 0.975 | 0.902 to 1.053 | 0.024 | 0.029 | 0.396 | 1.025 | 0.969 to 1.084 |
| Alcohol (log grams per day) | 1.092 | 0.571 | 0.056 | 2.980 | 0.972 to 9.134 | 1.449 | 0.739 | 0.050 | 4.258 | 1.001 to 18.119 | 3.506 | 0.501 | 2.53 x 10<sup>-12</sup> | 33.312 | 12.49 to 88.88 |
| Smoking (Ex vs Never) | 0.118 | 0.171 | 0.490 | 1.126 | 0.805 to 1.575 | 0.383 | 0.227 | 0.092 | 1.466 | 0.939 to 2.289 | 0.081 | 0.182 | 0.657 | 1.084 | 0.759 to 1.549 |
| Smoking (Current vs Never) | 1.063 | 0.170 | 3.66 x 10<sup>-10</sup> | 2.894 | 2.076 to 4.034 | 0.913 | 0.237 | 1.14 x 10<sup>-4</sup> | 2.491 | 1.567 to 3.960 | 1.072 | 0.179 | 2.18 x 10<sup>-9</sup> | 2.921 | 2.056 to 4.149 |
| Alcohol dependence | 0.609 | 0.203 | 0.0026 | 1.839 | 1.237 to 2.735 | 1.296 | 0.199 | 6.60 x 10<sup>-11</sup> | 3.655 | 2.477 to 5.394 | 2.027 | 0.134 | 2.28 x 10<sup>-51</sup> | 7.593 | 5.834 to 9.883 |
| Any alcoholic liver disease | 0.608 | 0.284 | 0.032 | 1.836 | 1.052 to 3.204 | 1.252 | 0.270 | 3.67 x 10<sup>-6</sup> | 3.498 | 2.058 to 5.963 | 2.092 | 0.163 | 9.99 x 10<sup>-38</sup> | 8.104 | 5.888 to 11.15 |
| *Multivariate analysis:* |  |  |  |  |  |  |  |  |  |  |  |  |  |  |  |
| Alcohol (log grams per day) | 0.733 | 0.573 | 0.201 | 2.081 | 0.676 to 6.402 | 0.645 | 0.759 | 0.396 | 1.905 | 0.430 to 8.440 | 2.272 | 0.513 | 9.29 x 10<sup>-6</sup> | 9.703 | 3.553 to 26.50 |
| Alcohol dependence | 0.395 | 0.206 | 0.055 | 1.485 | 0.992 to 2.224 | 1.152 | 0.207 | 2.65 x 10<sup>-8</sup> | 3.166 | 2.109 to 4.751 | 1.764 | 0.140 | 2.64 x 10<sup>-36</sup> | 5.834 | 4.433 to 7.679 |
| Smoking (Ex vs Never) | 0.124 | 0.171 | 0.468 | 1.133 | 0.809 to 1.585 | 0.405 | 0.227 | 0.075 | 1.500 | 0.961 to 2.342 | 0.134 | 0.182 | 0.463 | 1.143 | 0.800 to 1.633 |
| Smoking (Current vs Never) | 1.038 | 0.170 | 9.95 x 10<sup>-10</sup> | 2.825 | 2.024 to 3.941 | 0.825 | 0.238 | 5.19 x 10<sup>-4</sup> | 2.281 | 1.432 to 3.634 | 0.896 | 0.180 | 6.78 x 10<sup>-7</sup> | 2.450 | 1.720 to 3.488 |

### Biochemical and haematological risk factors for mortality in excessive drinkers

Results for many laboratory test results had substantial associations with overall survival (Table 5), particularly but not only the liver function tests. There were also strong but paradoxical associations with renal tests, with opposite effects for cystatin C (higher results = higher mortality) and urea or creatinine (lower results = higher mortality). Among the other biochemical tests, apolipoprotein B and LDL cholesterol were both negatively associated with mortality, as were IGF-1 and calcium. The association with calcium, which is partly bound to albumin in serum, became non-significant when albumin was also included in the survival analysis. C-reactive protein, sex-hormone-binding globulin, and testosterone (in men) were positively associated with all-cause mortality. Significant associations with all-cause mortality were also found for white cell count; red cell count, mean cell volume, and red cell distribution width; platelet count; reticulocyte maturity and size; and spherical cell volume.

**Table 5.** Laboratory test associations with all-cause mortality in excessive drinkers. Hazard Ratios (HR) are shown per 1-standard-deviation change in the age- and sex-adjusted standardised residuals for test results.

|  | Beta | SE | p-value | HR | 95% CI |
| --- | --- | --- | --- | --- | --- |
| *Liver tests:* |  |  |  |  |  |
| Alkaline phosphatase | 0.396 | 0.034 | $6.47 \times 10^{-32}$ | 1.486 | 1.391 to 1.587 |
| ALT | 0.052 | 0.037 | 0.165 | 1.053 | 0.979 to 1.134 |
| AST | 0.357 | 0.031 | $3.39 \times 10^{-31}$ | 1.430 | 1.346 to 1.518 |
| AST/ALT ratio | 0.325 | 0.028 | $1.90 \times 10^{-31}$ | 1.384 | 1.311 to 1.462 |
| GGT | 0.383 | 0.033 | $8.92 \times 10^{-31}$ | 1.466 | 1.374 to 1.564 |
| Total bilirubin | 0.056 | 0.037 | 0.136 | 1.057 | 0.983 to 1.137 |
| Direct bilirubin | 0.275 | 0.035 | $9.71 \times 10^{-15}$ | 1.316 | 1.228 to 1.411 |
| Indirect bilirubin | -0.010 | 0.039 | 0.804 | 0.990 | 0.918 to 1.069 |
| Total protein | 0.122 | 0.037 | 0.0011 | 1.130 | 1.050 to 1.216 |
| Albumin | -0.296 | 0.039 | $1.84 \times 10^{-14}$ | 0.744 | 0.690 to 0.802 |
| Globulins | 0.293 | 0.033 | $5.37 \times 10^{-19}$ | 1.341 | 1.257 to 1.430 |
| *Renal tests:* |  |  |  |  |  |
| Creatinine | -0.250 | 0.037 | $2.51 \times 10^{-11}$ | 0.779 | 0.724 to 0.838 |
| Cystatin C | 0.262 | 0.031 | $2.32 \times 10^{-17}$ | 1.300 | 1.223 to 1.381 |
| Urea | -0.456 | 0.040 | $6.39 \times 10^{-30}$ | 0.634 | 0.586 to 0.686 |
| Urate | -0.060 | 0.037 | 0.109 | 0.942 | 0.876 to 1.013 |
| *CVD-related tests:* |  |  |  |  |  |
| Glucose | 0.094 | 0.035 | 0.0073 | 1.098 | 1.026 to 1.177 |
| Glycated haemoglobin | 0.098 | 0.035 | 0.0056 | 1.103 | 1.029 to 1.181 |
| IGF-1 | -0.315 | 0.040 | $3.76 \times 10^{-15}$ | 0.730 | 0.675 to 0.789 |
| C-reactive protein | 0.218 | 0.037 | $2.40 \times 10^{-9}$ | 1.244 | 1.158 to 1.336 |
| Apo A | 0.052 | 0.038 | 0.170 | 1.054 | 0.978 to 1.135 |
| Apo B | -0.250 | 0.040 | $5.21 \times 10^{-10}$ | 0.779 | 0.719 to 0.843 |
| Lp(a) | -0.025 | 0.043 | 0.559 | 0.975 | 0.897 to 1.061 |
| LDL cholesterol | -0.264 | 0.039 | $2.19 \times 10^{-11}$ | 0.768 | 0.711 to 0.830 |
| HDL cholesterol | 0.117 | 0.035 | $8.20 \times 10^{-4}$ | 1.124 | 1.050 to 1.204 |
| Triglycerides | -0.111 | 0.039 | 0.0043 | 0.895 | 0.830 to 0.966 |
| *Bone-related tests:* |  |  |  |  |  |
| Calcium<sup>1</sup> | -0.149 | 0.039 | $1.19 \times 10^{-4}$ | 0.861 | 0.798 to 0.929 |
| Phosphate | 0.064 | 0.039 | 0.104 | 1.066 | 0.987 to 1.151 |
| Vitamin D | -0.295 | 0.044 | $1.73 \times 10^{-11}$ | 0.745 | 0.684 to 0.812 |
| *Endocrine tests:* |  |  |  |  |  |
| SHBG | 0.349 | 0.041 | $3.23 \times 10^{-17}$ | 1.417 | 1.307 to 1.537 |
| Oestradiol (M) | 0.043 | 0.074 | 0.563 | 1.044 | 0.903 to 1.207 |
| Oestradiol (F) | 0.245 | 0.272 | 0.368 | 1.277 | 0.750 to 2.175 |
| Testosterone (M) | 0.178 | 0.037 | $1.80 \times 10^{-6}$ | 1.195 | 1.111 to 1.285 |
| Testosterone (F) | 0.015 | 0.066 | 0.824 | 1.015 | 0.891 to 1.156 |
| *Haematology tests:* |  |  |  |  |  |
| White cell count | 0.175 | 0.035 | $7.33 \times 10^{-7}$ | 1.191 | 1.111 to 1.277 |
| Red cell count | -0.249 | 0.034 | $3.58 \times 10^{-13}$ | 0.780 | 0.729 to 0.834 |
| Haemoglobin | -0.022 | 0.037 | 0.547 | 0.978 | 0.911 to 1.051 |
| Haematocrit | -0.004 | 0.037 | 0.914 | 0.996 | 0.927 to 1.070 |
| MCV | 0.439 | 0.034 | $2.48 \times 10^{-38}$ | 1.551 | 1.451 to 1.658 |
| MCH | 0.322 | 0.029 | $7.77 \times 10^{-28}$ | 1.380 | 1.302 to 1.462 |
| MCHC | -0.046 | 0.036 | 0.203 | 0.955 | 0.889 to 1.025 |
| Red cell distribution width | 0.213 | 0.020 | $5.83 \times 10^{-27}$ | 1.237 | 1.190 to 1.286 |
| Platelet count | -0.012 | 0.038 | 0.755 | 0.988 | 0.918 to 1.064 |
| Platelet-Crit | -0.097 | 0.038 | 0.011 | 0.908 | 0.843 to 0.978 |
| Mean Platelet Volume | -0.101 | 0.038 | 0.0072 | 0.904 | 0.839 to 0.973 |
| Platelet Distribution Width | -0.046 | 0.037 | 0.213 | 0.955 | 0.888 to 1.027 |
| Lymphocyte count | -0.002 | 0.034 | 0.953 | 0.998 | 0.934 to 1.067 |
| Monocyte count | 0.179 | 0.030 | $1.27 \times 10^{-9}$ | 1.197 | 1.129 to 1.268 |
| Neutrophil count | 0.188 | 0.035 | $1.11 \times 10^{-7}$ | 1.207 | 1.126 to 1.294 |
| Eosinophil count | -0.040 | 0.038 | 0.291 | 0.961 | 0.892 to 1.035 |
| Basophil count | 0.078 | 0.034 | 0.022 | 1.081 | 1.011 to 1.156 |
| Reticulocyte count | -0.006 | 0.038 | 0.866 | 0.994 | 0.921 to 1.071 |
| Immature Reticulocytes | 0.135 | 0.037 | $2.29 \times 10^{-4}$ | 1.145 | 1.065 to 1.230 |
| Mean Reticulocyte Volume | 0.372 | 0.034 | $2.28 \times 10^{-28}$ | 1.451 | 1.358 to 1.550 |
| Mean Spherical Cell Volume | 0.457 | 0.033 | $5.83 \times 10^{-44}$ | 1.579 | 1.481 to 1.684 |

<sup>1</sup> Calcium results are not adjusted for albumin concentration

When causes of death were divided into cancers, circulatory diseases and other causes (see Supplementary Table 5) a number of differences were seen. For cancers, higher values for the liver function tests alkaline phosphatase and GGT, and for C-reactive protein, were associated with greater mortality. On the other hand higher values for urea and vitamin D were associated with lower cancer mortality. Among the haematology results, measures related to erythrocyte and reticulocyte size (MCV, RDW, reticulocyte and spherical cell volumes) were significantly related to risk of death from any cancer.

For deaths due to circulatory diseases, many of the liver function tests showed positive (higher result = higher risk) associations. These included the liver enzymes alkaline phosphatase, AST (but not ALT) and GGT, direct (conjugated) bilirubin and globulins. Albumin showed a negative association. In addition the renal tests urea and cystatin C showed significant associations, though in opposite directions, and higher values for both apolipoprotein B and LDL cholesterol were associated with lower risk. The haematology tests associated with cardiovascular deaths were similar to those identified for cancer deaths; size of erythrocytes and related cells in the blood.

For deaths from all other causes except liver disease, the liver function and renal tests were significantly associated with risk and in addition there were associations with IGF-1, C-reactive protein, apolipoprotein B and both LDL and HDL cholesterol, calcium, vitamin D, SHBG and testosterone (in men). The haematology tests showing significant association with other-cause deaths included the erythrocyte-size-related ones mentioned previously, and also the red and white cell (total, monocyte and neutrophil) counts.

## DISCUSSION

As expected, the high and persistent alcohol intake which defined the excessive drinker group was associated with increased mortality compared to other UK Biobank participants. However many of the people in this category do survive into old age, and apart from an excess of liver disease the causes of death are similar to those for other participants. Within the excessive-drinking group, risk factors with substantial effects on mortality have been identified; firstly alcohol dependence, and secondly liver disease (either as a documented clinical diagnosis or inferred from abnormality in biochemical liver function tests). Smoking, which is common among the excessive drinkers, also affects mortality within this group.

### Mortality associated with excessive drinking

All-cause mortality was substantially increased in UK Biobank participants who reported long-term very high alcohol intake, with all-cause mortality risk (measured as hazard ratio) approximately two-fold compared to all other participants. HRs were similar in the men and women despite the different cut-offs for excessive drinking, supporting the decision to use 80 g/day for men and 50 g/day for women. This two-fold HR estimate is based on a reference group comprising everyone who did not meet the definition of excessive drinker, and when a more tightly defined reference group consisting of those who reported some but low-risk alcohol intake (5-15 grams/day) was used instead then the all-cause mortality HR was slightly higher at 2.5 (Supplementary Table 2). In contrast, the liver disease mortality HR estimate was doubled from 9-fold for the excessive drinkers versus all others comparison, to 18-fold when the reference group was the 5-15 g/d drinkers. The increases in risk difference may be due to exclusion of current non-drinkers, or exclusion of those reporting over 15 g/d but not meeting the excessive drinker criteria, or both.

Inspection of the proportions for underlying or primary causes of death, coded as ICD-10 diagnoses, showed that cancers (C00-C99) and circulatory diseases (I00-I99) were the most common categories in both the excessive drinkers and all other participants (see Figure 2, panel A). Out of the major ICD-10 categories, only diseases of the digestive system (K00-K93, which includes liver diseases as K70-K76) comprised a significantly higher proportion of deaths in the excessive drinkers than in all others. Survival analysis by reported cause of death (Figure 2, panel B), showed HRs significantly above 1 for most groups of causes and with confidence intervals which included the all-cause mortality HR of 2.0. So although excessive drinkers have poorer survival (HR > 1), the distribution of causes of death other than liver disease is similar to that for the rest of the population.

Smoking, which is more common among the excessive drinkers than among the other participants, accounts for part of the mortality difference between these groups. It is worth pointing out that although mortality is higher in the excessive drinkers, not all of this increase is directly attributable to alcohol. At least part is due to other characteristics of the excessive drinker group. Most large-scale studies on alcohol-related mortality have accounted for smoking in their data analysis, so comparisons between our HR estimates and previous ones should take this into account (relevant information is shown in Supplementary Table 4).

### Risk factors for mortality among excessive drinkers

Although survival curves appear to diverge by age 60 (Figure 1), around three-quarters of the male excessive drinkers are projected to live to age 78 and of the female excessive drinkers, to 81. There is substantial variation in mortality within the excessive drinking group, and we have identified multiple factors associated with differences in risk.

Conventional risk factors such as obesity and blood pressures were not good predictors of mortality in the excessive drinkers. This may be a matter of power (numbers and length of follow-up) but is quite likely due to other factors taking a major role. Given the association between obesity and liver disease, including alcohol-related liver disease, the lack of association between BMI or WHR and mortality is surprising but may be due to poor nutrition contributing to poor outcomes.

As can be seen from Supplementary Figures 1 and 2, the effects of alcohol dependence and alcohol-related liver disease are greater than those of excessive drinking alone on survival. The importance of alcohol dependence rather than quantitative alcohol intake supports conclusions from previous studies (14-16), in which mortality associations with symptom count for alcohol dependence, or with alcohol dependence diagnosis, were more significant than those with the participants’ reported alcohol intake. Alcohol dependence and high alcohol intake are strongly associated but have at least some differences in their causes, as shown by the genetic correlation between them being significantly less than 1 (17, 18). In this analysis of mortality affecting excessive drinkers, the presence of a recorded diagnosis of alcohol dependence had a substantial and detrimental association with survival which applied across the major groupings of causes of death (except for cancers, for which p = 0.055).

Our results cannot answer the question of how alcohol dependence (as distinct from high alcohol intake) leads to or is associated with increased mortality. One obvious possibility is that those who are dependent consume more alcohol, but the multivariate analysis including both dependence and intake does not support this. Another hypothesis is that those with dependence are likely to have poorer social support and poorer nutrition than people who drink excessively without diagnosed dependence. Alcohol dependence is likely to co-exist with other substance use disorders or psychiatric conditions, and possibly with despair and feelings of being unable to control events, but the biological link between these and mortality is unclear.

Many biochemical and metabolic factors are known to be affected by alcohol intake (19), and several of these, particularly liver enzymes, are associated with mortality in the general population (20). Although it is reasonable to suppose that abnormal liver function tests in excessive drinkers without known liver disease are in fact an indication of liver damage, and will be associated with a poorer prognosis, there is little published information to substantiate this. Here we show that measures of liver damage (alkaline phosphatase, AST, GGT), and hepatic excretory and synthetic function (direct bilirubin, albumin) significantly predict deaths from any cause (Table 5), from cancers, cardiovascular disease (Supplementary Table 5), and from liver disease (Table 2). Having results in the top quintile for alkaline phosphatase, AST or GGT is associated with a more than two-fold risk for all-cause mortality compared to those in the lowest quintile for excessive drinkers (see Supplementary Figure 3). However mortality was not associated with ALT except for deaths ascribed to liver disease, and the AST/ALT ratio offered no predictive advantage over AST alone.

Direct bilirubin was associated with mortality but total and indirect bilirubin were not. The probable explanation is that the ‘signal’ (direct bilirubin being associated with mortality) is obscured by ‘noise’ from variation in indirect bilirubin (which is not associated) when total bilirubin is tested as the predictor variable. Indirect or unconjugated bilirubin, mainly reflecting red cell turnover, makes up about four-fifths of the total bilirubin in both the overall UK Biobank population and in the excessive drinkers. Direct bilirubin reflects defective excretion after conjugation of bilirubin in the liver, and is therefore a more sensitive index of minor or early alcohol-related liver dysfunction.

Among the excessive drinkers in UK Biobank, liver function tests and presence of liver disease are associated with causes of death not obviously involving the liver, particularly cardiovascular disease. A study in Sweden showed that alcohol-related liver disease was associated with significantly (> 4-fold) increased all-cause mortality, with >40% of these deaths being liver-related (21). The association between liver function tests and all-cause or cardiovascular mortality is well-documented for the general population (20, 22, 23), consistent with reports associating non-alcoholic liver disease or liver enzymes with cardiovascular disease risk (24, 25).

Several measures related to erythrocyte size, including red cell count, mean cell volume, red cell distribution width, and reticulocyte and spherical cell size, were strongly associated with mortality (larger cells = higher risk) in the excessive drinkers. Examination of the correlation matrix, and factor analysis of these tests (results not shown), suggested that all except red cell distribution width were associated with each other and presumably with variation in erythrocyte maturation and/or red cell membrane characteristics. Erythrocyte size (MCV) has long been recognised as being increased by high alcohol intake, and a small prospective study in patients attending a hospital emergency department (26) showed increased rates of alcohol-related illness including liver disease or gastrointestinal bleeding in men in the top quintile for MCV. Association between erythrocyte size and mortality, including mortality from primary liver cancer has also been reported (27) in the general population,. Red cell distribution width has been shown to be associated with mortality, but most reports are on patients with renal disease (28) or on acute mortality after hospital admission (29, 30). It is currently unknown how variation in erythrocyte characteristics in excessive drinkers (or indeed in others) is associated with variation in mortality.

Other test results associated with all-cause mortality include markers of renal function. Both urea and creatinine showed inverse associations (higher result = lower risk) with mortality. It is possible that the association with urea is due to impairment of liver function, as urea is synthesised in the liver, but this does not apply to creatinine and it seems probable that the associations for these two tests are due to them acting as markers of protein intake and muscle mass. On the other hand the associations for cystatin C, another measure of glomerular filtration rate, are positive (higher result = higher risk). Cystatin C is derived from nucleated cells, is cleared from the blood by the kidneys, and is less subject to the non-renal sources of variation affecting urea or creatinine. It appears that mortality in excessive drinkers can be affected by both decreased renal function and poor nutritional status.

Several other tests showed significant associations with mortality which were consistent with those reported for the general population. C-reactive protein reflects chronic inflammation and predicts all-cause and cardiovascular mortality (31), and the similar association in excessive drinkers is to be expected. Lower vitamin D results were associated with higher mortality, again as expected from previous studies (32). The associations for IGF-1, SHBG and testosterone, although strong and consistent with at least some previous reports (33, 34), do not have ready explanations.

Lipid risk factors such as apolipoproteins and lipoprotein cholesterol fractions had associations with mortality which were unlike those expected for the general population. Higher levels of apolipoprotein B and LDL cholesterol were associated with lower mortality from all causes, from cardiovascular diseases and from liver diseases, and the association was in the same direction for cancers. As discussed for urea and creatinine above, this may be due to poor nutrition in some of the excessive drinkers affecting the test results and decreasing survival. Alternatively or additionally, patients with liver disease (which is associated with poorer prognosis) may be less able to synthesise low-density lipoproteins.

### Limitations

The nature of the data used for this analysis imposes some limitations. Only those who survived to age 40 were eligible for participation in the UK Biobank, so alcohol-associated mortality earlier in life could not be assessed. The conclusions may not be applicable to non-UK populations, although risk factor information can probably be generalised to other economically advanced societies. Only a small proportion of participants had died by the cut-off date used; this does not appear to lead to insufficient power because the starting number was large but the causes of death may change over time as the average age of the cohort increases. Some of those who did not meet both the current and ten-years-previously criteria for excessive drinking would have been drinking at hazardous levels, or may have done so previously. This will tend to diminish rather than increase the mortality differences between the excessive drinkers and others and it will not have affected the identification of factors influencing risk within the excessive-drinker group. Diagnoses may be incomplete, with disease being present but not recorded during life; and causes of death listed on death certificates can be inaccurate. In relation to the two risk factors of diagnosed alcohol dependence and alcohol-related liver disease, it is possible these overlap because identification of alcohol dependence may have led to diagnosis of alcohol-related liver disease, or *vice versa*.

Overall, the strengths afforded by such a large unselected cohort outweigh the limitations, and make it possible to address questions about the natural history of high alcohol intake in ways which have not previously been possible.

### Clinical and research implications

Clinically, identification and early intervention for excessive drinkers with alcohol dependence and/or alcohol-related liver damage could divert at least some of them towards abstinence, and there is evidence (35) that feedback about liver damage can motivate patients to reduce alcohol intake. For research directions, alcohol-related liver disease risk in excessive drinkers is known to be affected by genetic variation (36), and larger studies should provide additional loci, define likely pathways, and lead to investigation of possible associations between polygenic risk scores for liver disease and mortality. Similarly, identification of causes of alcohol dependence, distinct from those affecting alcohol intake, would allow testing of these against the mortality data. The role of alcohol in changing erythrocyte and erythrocyte precursor size and membrane characteristics, and how these affect mortality, also deserves further investigation.

## Supporting information

STROBE Checklist

## Data Availability

All data used in this study are from the UK Biobank, which can grant access to approved researchers.

## ABBREVIATIONS

ALT: Alanine aminotransferase
AST: Aspartate aminotransferase
BMI: Body Mass Index
CI: Confidence Interval
DBP: Diastolic Blood Pressure
GGT: Gamma-glutamyl transferase
HDL: High-density lipoprotein
HR: Hazard Ratio
ICD-10: International Classification of Diseases, 10^th^ Edition
IGF-1: Insulin-like growth factor 1
LDL: Low-density lipoprotein
Lp(a): Lipoprotein (a)
MCV: Mean cell volume (of erythrocytes)
MRV: Mean reticulocyte volume
MSCV: Mean spherical cell volume
RDW: Red cell distribution width
SBP: Systolic Blood Pressure
SE: Standard Error
SHBG: Sex-hormone-binding globulin
WHR: Waist/Hip Ratio

## Appendix 1. List of Investigators, GenomALC Consortium

Guruprasad P. Aithal PhD FRCP NIHR Nottingham Biomedical Research Centre, Nottingham University Hospitals and the University of Nottingham, Nottingham NG7 2UH, United Kingdom

Stephen R. Atkinson PhD Department of Metabolism, Digestion and Reproduction, Imperial College London, London, United Kingdom

Ramon Bataller MD PhD Center for Liver Diseases, University of Pittsburgh Medical Center, 3471 Fifth Avenue, Pittsburgh, PA 15213, USA

Gregory Botwin PhD Translational Genomics Group, Inflammatory Bowel & Immunobiology Research Institute, 8700 Beverly Blvd., Thalians Bldg. #E244 : Los Angeles CA 90048. F. Widjaja Family Foundation Inflammatory Bowel and Immunobiology Research Institute, Cedars-Sinai Medical Center, Los Angeles, California, CA 90822, USA

Naga P. Chalasani MD Department of Medicine, Indiana University, Indianapolis, IN 46202-5175, USA

Heather J. Cordell DPhil Population Health Sciences Institute, Faculty of Medical Sciences, Newcastle University, International Centre for Life, Central Parkway, Newcastle upon Tyne NE1 3BZ, United Kingdom

Ann K. Daly PhD Faculty of Medical Sciences, Newcastle University Medical School, Framlington Place, Newcastle upon Tyne NE2 4HH, United Kingdom

Rebecca Darlay PhD Population Health Sciences Institute, Faculty of Medical Sciences, Newcastle University, International Centre for Life, Central Parkway, Newcastle upon Tyne NE1 3BZ, United Kingdom

Christopher P. Day MD, PhD Newcastle University, Framlington Place, Newcastle upon Tyne NE2 4HH, United Kingdom

Florian Eyer MD Division of Clinical Toxicology, Department of Internal Medicine 2, Klinikum rechts der Isar, School of Medicine, Technical University of Munich, Ismaninger Str. 22, 81675 Munich, Germany

Tatiana Foroud PhD Department of Medical and Molecular Genetics, Indiana University, USA

Dermot Gleeson MD FRCP Liver Unit, Sheffield Teaching Hospitals, AO Floor Robert Hadfield Building, Northern General Hospital, Sheffied S5 7AU, UK.

David Goldman MD Laboratory of Neurogenetics, NIAAA, Rockville, MD 20852, USA

Paul S. Haber MD PhD Drug Health Services, Royal Prince Alfred Hospital, Missenden Road, Camperdown, NSW 2050, Australia Faculty of Medicine and Health, The University of Sydney, Sydney, NSW 2006, Australia

Jean-Marc Jacquet MD Service Addictologie, CHRU Caremeau, 30029 Nîmes, France

Tiebing Liang PhD Department of Medicine, Indiana University, Indianapolis, IN 46202-5175, USA

Suthat Liangpunsakul MD Division of Gastroenterology and Hepatology, Department of Medicine, Indiana University, Indianapolis and and Roudebush Veterans Administration Medical Center, Indianapolis, IN, USA

Steven Masson FRCP Faculty of Medical Sciences, Newcastle University Medical School, Framlington Place, Newcastle upon Tyne NE2 4HH, United Kingdom

Philippe Mathurin MD PhD CHRU de Lille, Hôpital Claude Huriez, Rue M. Polonovski CS 70001, 59 037 Lille Cedex, France

Romain Moirand MD PhD Univ Rennes, INRA, INSERM, CHU Rennes, Institut NUMECAN (Nutrition Metabolisms and Cancer), F-35000 Rennes, France

Christophe Moreno MD PhD CUB Hôpital Erasme, Université Libre de Bruxelles, Brussels, Belgium

Timothy R. Morgan MD Department of Veterans Affairs, VA Long Beach Healthcare System, 5901 East Seventh Street, Long Beach, CA 90822, USA Department of Medicine, University of California, Irvine, USA

Marsha Morgan MD PhD UCL Institute for Liver & Digestive Health, Division of Medicine, Royal Free Campus, University College London, London NW3 2PF, UK

Sebastian Mueller MD PhD Department of Internal Medicine, Salem Medical Center and Center for Alcohol Research, University of Heidelberg, Zeppelinstraße 11-33, 69121 Heidelberg, Germany

Beat Müllhaupt MD Department of Gastroenterology and Hepatology, University Hospital Zurich, Rämistrasse 100, CH-8901 Zurich, Switzerland

Laura E. Nagy PhD Lerner Research Institute, 9500 Euclid Avenue, Cleveland, Ohio, OH 44195, USA

Pierre Nahon MD PhD AP-HP, Hôpital Avicenne, Liver Unit, Bobigny, France Inserm U1162 Génomique fonctionnelle des tumeurs solides, Paris, France

Bertrand Nalpas MD PhD Service Addictologie, CHRU Caremeau, 30029 Nîmes, France DISC, Inserm, 75013 Paris, France

Sylvie Naveau MD Hôpital Antoine-Béclère, 157 Rue de la Porte de Trivaux, 92140 Clamart, France

Pascal Perney MD PhD Hôpital Universitaire Caremeau, Place du Pr. Robert Debre, 30029 Nîmes, France, UM1, INSERM U1018

Munir Pirmohamed PhD FRCP MRC Centre for Drug Safety Science and Wolfson Centre for Personalised Medicine, Institute of Translational Medicine, University of Liverpool, Liverpool, L69 3GL, UK MRC Centre for Drug Safety Science, Liverpool Centre for Alcohol Research, University of Liverpool, The Royal Liverpool and Broadgreen University Hospitals NHS Trust, and Liverpool Health Partners, Liverpool, L69 3GL, UK

Tae-Hwi Schwantes-An PhD Department of Medical and Molecular Genetics, Indiana University, USA

Helmut K. Seitz MD Department of Internal Medicine, Salem Medical Center and Center for Alcohol Research, University of Heidelberg, Zeppelinstraße 11e33, 69121 Heidelberg, Germany

Devanshi Seth PhD Edith Collins Centre (Translational Research in Alcohol Drugs and Toxicology), Drug Health Services, Sydney Local Health District, KGV Building, 83-117 Missenden Road, Camperdown, NSW 2050, Australia Centenary Institute of Cancer Medicine and Cell Biology, The University of Sydney, Sydney, NSW 2006, Australia Faculty of Medicine and Health, The University of Sydney, Sydney, NSW 2006, Australia

Michael Soyka MD; Psychiatric Hospital University of Munich, Nussbaumsstr.7, 80336 Munich, Germany and Privatklinik Meiringen, Willigen, CH 3860 Meiringen, Switzerland

Felix Stickel MD PhD Department of Gastroenterology and Hepatology, University Hospital Zurich, Rämistrasse 100, CH-8901 Zurich, Switzerland

Andrew Thompson PhD Health Analytics, Lane Clark & Peacock LLP, London, UK MRC Centre for Drug Safety Science, Liverpool Centre for Alcohol Research, University of Liverpool, The Royal Liverpool and Broadgreen University Hospitals NHS Trust, and Liverpool Health Partners, Liverpool, L69 3GL, UK

Mark R. Thursz MD Institute for Liver & Digestive Health, Imperial College, London, UK

Eric Trepo MD PhD Université Libre de Bruxelles, Brussels, Belgium

John B. Whitfield PhD FRCPath Genetic Epidemiology, QIMR Berghofer Medical Research Institute, Queensland 4029, Australia

## SUPPLEMENTARY MATERIAL

**Supplementary Table 1.** Descriptive statistics (percentages, or medians, 20^th^ and 80^th^ centiles, and numbers of people with data) for UK Biobank participants at the time of initial assessment (except for alcohol dependence, which is based on hospital or death certificate data). Presence of diabetes is based on whether the participant had been told by a doctor that they had diabetes.

|  |  | **Excessive drinkers (N=6640)** |  | **All others (N=495,866)** |  |
| --- | --- | --- | --- | --- | --- |
|  |  | **Men (N=5136)** | **Women (N=1504)** | **Men (N=223,985)** | **Women (N=271,879)** |
| Age at assessment (years) |  | 57 (49, 64) 5136 | 55 (47, 62) 1504 | 58 (48, 65) 223,985 | 57 (48, 64) 271,879 |
| BMI (kg/m<sup>2</sup>) |  | 28.0 (24.7, 31.6) 5105 | 26.0 (23.1, 30.5) 1496 | 27.3 (24.4, 30.8) 222,370 | 26.1 (22.9, 30.8) 270,428 |
| Systolic Blood Pressure |  | 147 (133, 162) 4817 | 138 (122, 155) 1428 | 139 (126,155) 210,725 | 133 (119, 151) 255,415 |
| Diastolic Blood Pressure |  | 88 (79, 96) 4817 | 84 (75, 92) 1428 | 84 (76, 92) 210,727 | 80 (72, 89) 255,418 |
| Smoking: | Never | 1300 (25.4%) | 377 (25.1%) | 110148 (49.5%) | 161648 (59.8%) |
|  | Former | 2268 (44.3%) | 684 (45.6%) | 85340 (38.3%) | 84766 (31.4%) |
|  | Current | 1556 (30.4%) | 439 (29.3%) | 27055 (12.2%) | 23928 (8.9%) |
| Diabetes |  | 314/5112 (6.1%) | 39/1502 (2.6%) | 15,686/222,596 (7.0%) | 10,360/270,679 (3.8%) |
| Alcohol intake (grams/day) |  | 95 (85, 119) 5136 | 60 (52, 75) 1504 | 16.2 (1.2, 35.7) 233,985 | 6.3 (0, 16.7) 271,879 |
| Alcohol dependence |  | 346/5136 (6.7%) | 40/1504 (2.7%) | 1681/223,986 (0.8%) | 588/271,878 (0.2%) |
| Albumin (g/l) |  | 45.4 (43.2, 47.6) 4430 | 45.3 (43.3, 47.6) 1281 | 45.5 (43.4, 47.7) 194,178 | 44.9 (42.8, 47.1) 230,187 |
| Bilirubin (total) (μmol/l) |  | 9.1 (6.9, 12.4) 4775 | 7.7 (5.8, 10.2) 1391 | 9.1 (7.0, 12.5) 209,246 | 7.3 (5.6, 9.8) 252,149 |
| ALP (units/l) |  | 81 (66, 102) 4804 | 79 (62, 101) 1400 | 79 (64, 97) 210,160 | 82 (64, 103) 253,239 |
| AST (units/l) |  | 29 (24, 39) 4774 | 24 (20, 30) 1391 | 26 (22, 32) 209,315 | 23 (19, 28) 252,317 |
| ALT (units/l) |  | 27 (19, 41) 4800 | 19 (13, 28) 1400 | 24 (17, 35) 209,990 | 18 (13, 25) 253,212 |
| GGT (units/l) |  | 62 (35, 124) 4791 | 31 (20, 57) 1397 | 33 (22, 55) 210,040 | 21 (15, 36) 253,115 |
| Mean cell volume (fl) |  | 94.5 (90.9, 98.4) 4890 | 94.6 (91.2, 98.2) 1425 | 91.3 (88.1, 94.7) 214,344 | 91.1 (87.6, 94.4) 257,460 |

**Supplementary Table 2.**
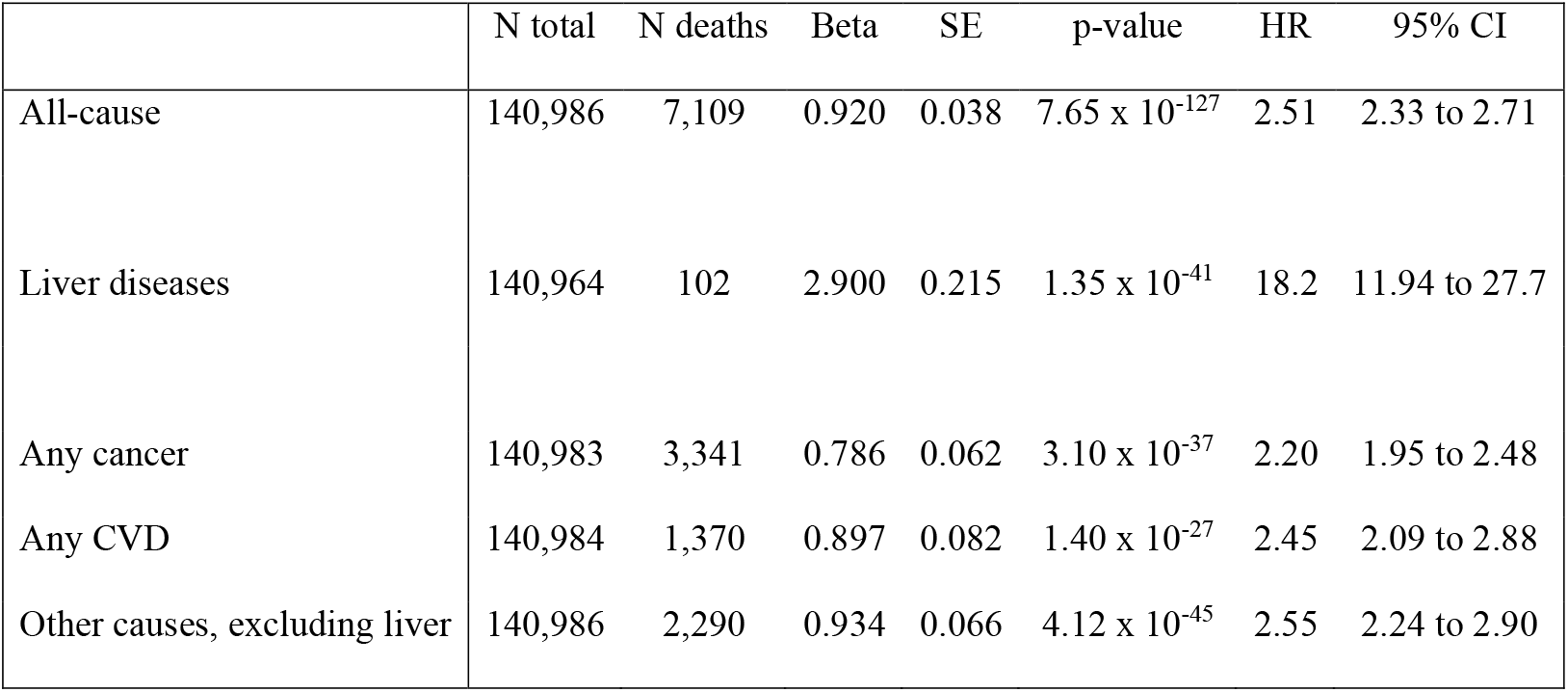
Survival analysis by Cox regression comparing excessive drinkers versus participants reporting alcohol intake of 5-15 grams/day. In each case sex has been included as an additional factor affecting survival. All-cause mortality, and mortality from any cancer, any cardiovascular disease, all other causes except liver diseases, and liver disease. SE, standard error; HR, Hazard Ratio; 95% CI, 95% confidence interval for the Hazard Ratios.

**Supplementary Table 3.**
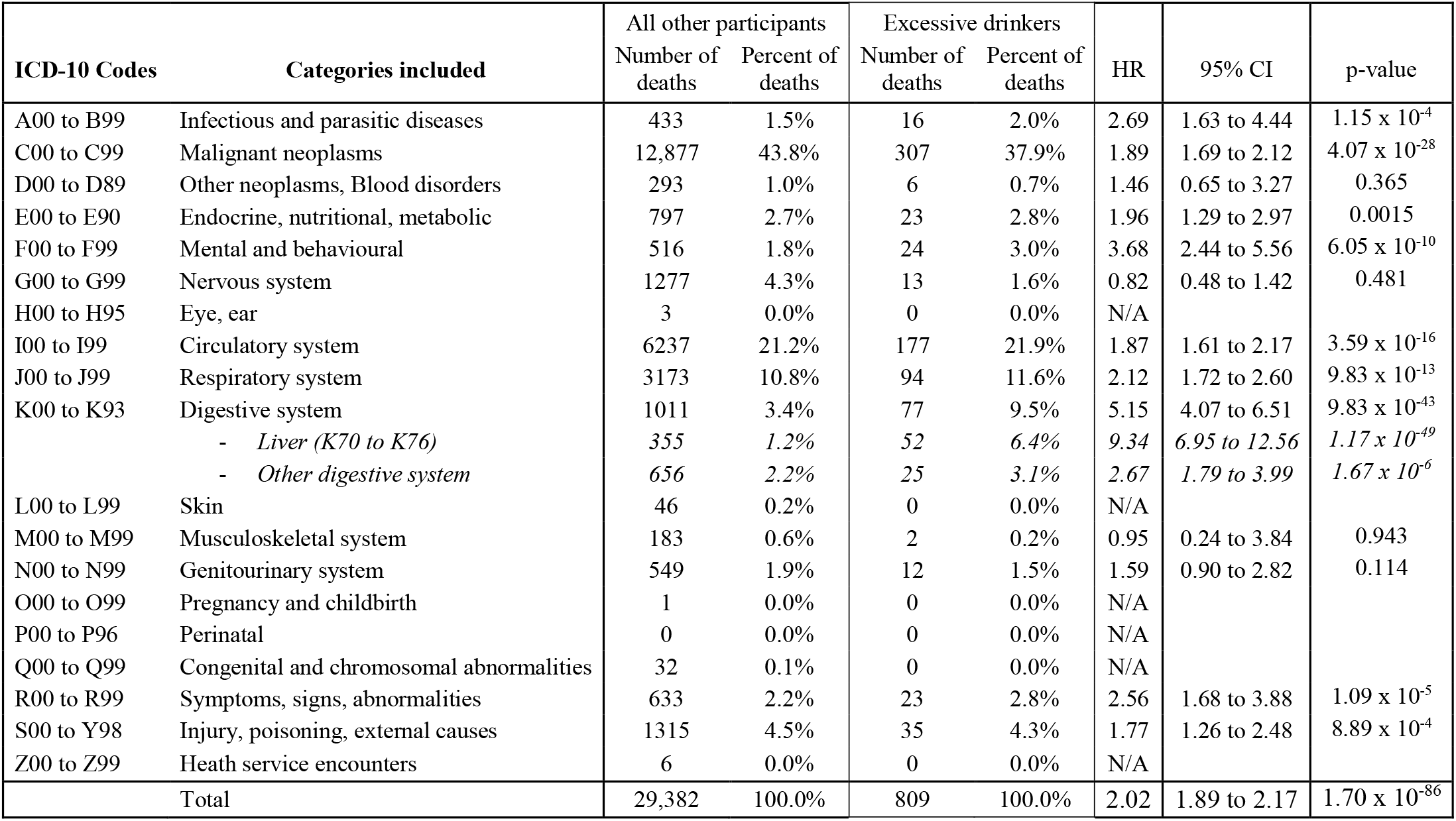
Comparison of reported causes of death between excessive drinkers and all other participants. The HR estimates and p-values are from survival analysis rather than a direct comparison of proportions.

| ICD-10 Codes | Categories included | All other participants |  | Excessive drinkers |  | HR | 95% CI | p-value |
| --- | --- | --- | --- | --- | --- | --- | --- | --- |
|  |  | Number of deaths | Percent of deaths | Number of deaths | Percent of deaths |  |  |  |
| A00 to B99 | Infectious and parasitic diseases | 433 | 1.5% | 16 | 2.0% | 2.69 | 1.63 to 4.44 | $1.15 \times 10^{-4}$ |
| C00 to C99 | Malignant neoplasms | 12,877 | 43.8% | 307 | 37.9% | 1.89 | 1.69 to 2.12 | $4.07 \times 10^{-28}$ |
| D00 to D89 | Other neoplasms, Blood disorders | 293 | 1.0% | 6 | 0.7% | 1.46 | 0.65 to 3.27 | 0.365 |
| E00 to E90 | Endocrine, nutritional, metabolic | 797 | 2.7% | 23 | 2.8% | 1.96 | 1.29 to 2.97 | 0.0015 |
| F00 to F99 | Mental and behavioural | 516 | 1.8% | 24 | 3.0% | 3.68 | 2.44 to 5.56 | $6.05 \times 10^{-10}$ |
| G00 to G99 | Nervous system | 1277 | 4.3% | 13 | 1.6% | 0.82 | 0.48 to 1.42 | 0.481 |
| H00 to H95 | Eye, ear | 3 | 0.0% | 0 | 0.0% | N/A |  |  |
| I00 to I99 | Circulatory system | 6237 | 21.2% | 177 | 21.9% | 1.87 | 1.61 to 2.17 | $3.59 \times 10^{-16}$ |
| J00 to J99 | Respiratory system | 3173 | 10.8% | 94 | 11.6% | 2.12 | 1.72 to 2.60 | $9.83 \times 10^{-13}$ |
| K00 to K93 | Digestive system | 1011 | 3.4% | 77 | 9.5% | 5.15 | 4.07 to 6.51 | $9.83 \times 10^{-43}$ |
| | - Liver (K70 to K76) | 355 | 1.2% | 52 | 6.4% | 9.34 | 6.95 to 12.56 | $1.17 \times 10^{-49}$ |
| | - Other digestive system | 656 | 2.2% | 25 | 3.1% | 2.67 | 1.79 to 3.99 | $1.67 \times 10^{-6}$ |
| L00 to L99 | Skin | 46 | 0.2% | 0 | 0.0% | N/A |  |  |
| M00 to M99 | Musculoskeletal system | 183 | 0.6% | 2 | 0.2% | 0.95 | 0.24 to 3.84 | 0.943 |
| N00 to N99 | Genitourinary system | 549 | 1.9% | 12 | 1.5% | 1.59 | 0.90 to 2.82 | 0.114 |
| O00 to O99 | Pregnancy and childbirth | 1 | 0.0% | 0 | 0.0% | N/A |  |  |
| P00 to P96 | Perinatal | 0 | 0.0% | 0 | 0.0% | N/A |  |  |
| Q00 to Q99 | Congenital and chromosomal abnormalities | 32 | 0.1% | 0 | 0.0% | N/A |  |  |
| R00 to R99 | Symptoms, signs, abnormalities | 633 | 2.2% | 23 | 2.8% | 2.56 | 1.68 to 3.88 | $1.09 \times 10^{-5}$ |
| S00 to Y98 | Injury, poisoning, external causes | 1315 | 4.5% | 35 | 4.3% | 1.77 | 1.26 to 2.48 | $8.89 \times 10^{-4}$ |
| Z00 to Z99 | Health service encounters | 6 | 0.0% | 0 | 0.0% | N/A |  |  |
| Total | | 29,382 | 100.0% | 809 | 100.0% | 2.02 | 1.89 to 2.17 | $1.70 \times 10^{-86}$ |

**Supplementary Table 4.** Comparison of Hazard Ratios between excessive drinkers and all other participants, showing results adjusted for sex only, then with addition of information on smoking status, then with addition of information on other recognised risk factors (as listed in the footnote).

|  | Sex only |  |  | Add smoking |  |  | Add others* |  |  |
| --- | --- | --- | --- | --- | --- | --- | --- | --- | --- |
|  | HR | 95% CI | p-value | HR | 95% CI | p-value | HR | 95% CI | p-value |
| All-cause | 2.02 | 1.89 to 2.17 | $1.70 \times 10^{-86}$ | 1.66 | 1.54 to 1.78 | $7.77 \times 10^{-45}$ | 1.70 | 1.56 to 1.84 | $9.86 \times 10^{-37}$ |
| Infectious and parasitic diseases | 2.69 | 1.63 to 4.44 | $1.15 \times 10^{-4}$ | 2.26 | 1.36 to 3.74 | 0.0016 | 2.29 | 1.24 to 4.22 | 0.0081 |
| Malignant neoplasms | 1.89 | 1.69 to 2.12 | $4.07 \times 10^{-28}$ | 1.58 | 1.41 to 1.78 | $2.97 \times 10^{-15}$ | 1.65 | 1.45 to 1.88 | $8.52 \times 10^{-14}$ |
| Other neoplasms, Blood disorders | 1.45 | 0.65 to 3.27 | 0.365 | 1.37 | 0.61 to 3.09 | 0.450 | 1.65 | 0.67 to 4.06 | 0.273 |
| Endocrine, nutritional, metabolic | 1.96 | 1.29 to 2.97 | 0.0015 | 1.68 | 1.11 to 2.56 | 0.015 | 1.75 | 1.06 to 2.89 | 0.029 |
| Mental and behavioural | 3.68 | 2.44 to 5.56 | $6.05 \times 10^{-10}$ | 3.21 | 2.12 to 4.87 | $3.88 \times 10^{-8}$ | 2.86 | 1.74 to 4.70 | $3.25 \times 10^{-5}$ |
| Nervous system | 0.82 | 0.47 to 1.42 | 0.481 | 0.81 | 0.47 to 1.40 | 0.447 | 0.73 | 0.36 to 1.47 | 0.374 |
| Circulatory system | 1.87 | 1.61 to 2.17 | $3.59 \times 10^{-16}$ | 1.49 | 1.28 to 1.73 | $2.82 \times 10^{-7}$ | 1.50 | 1.26 to 1.78 | $5.45 \times 10^{-6}$ |
| Respiratory system | 2.12 | 1.72 to 2.60 | $9.83 \times 10^{-13}$ | 1.55 | 1.26 to 1.91 | $2.97 \times 10^{-5}$ | 1.53 | 1.21 to 1.95 | $4.46 \times 10^{-4}$ |
| Digestive system | 5.15 | 4.07 to 6.51 | $7.48 \times 10^{-43}$ | 3.98 | 3.14 to 5.05 | $3.01 \times 10^{-30}$ | 3.74 | 2.81 to 4.97 | $1.03 \times 10^{-19}$ |
| Liver | 9.34 | 6.95 to 12.56 | $1.17 \times 10^{-49}$ | 7.25 | 5.36 to 9.80 | $8.87 \times 10^{-38}$ | 6.67 | 4.63 to 9.62 | $2.72 \times 10^{-24}$ |
| Other | 2.67 | 1.79 to 3.99 | $1.67 \times 10^{-6}$ | 2.06 | 1.38 to 3.09 | $4.46 \times 10^{-4}$ | 2.01 | 1.24 to 3.24 | 0.0043 |
| Musculoskeletal system | 0.95 | 0.24 to 3.84 | 0.943 | 0.82 | 0.20 to 3.34 | 0.785 | 1.24 | 0.30 to 5.09 | 0.762 |
| Genitourinary system | 1.59 | 0.89 to 2.82 | 0.114 | 1.34 | 0.76 to 2.39 | 0.313 | 1.61 | 0.90 to 2.88 | 0.107 |
| Symptoms, signs, abnormalities | 2.56 | 1.68 to 3.88 | $1.09 \times 10^{-5}$ | 2.06 | 1.35 to 3.14 | $7.73 \times 10^{-4}$ | 2.57 | 1.61 to 4.10 | $7.99 \times 10^{-5}$ |
| Injury, poisoning, external causes | 1.77 | 1.26 to 2.48 | $8.89 \times 10^{-4}$ | 1.46 | 1.04 to 2.05 | 0.029 | 1.42 | 0.95 to 2.12 | 0.089 |

\* Additional risk factors are BMI, WHR, age completed full-time education, self-reported diabetes, systolic and diastolic blood pressures.

**Supplementary Table 5.**
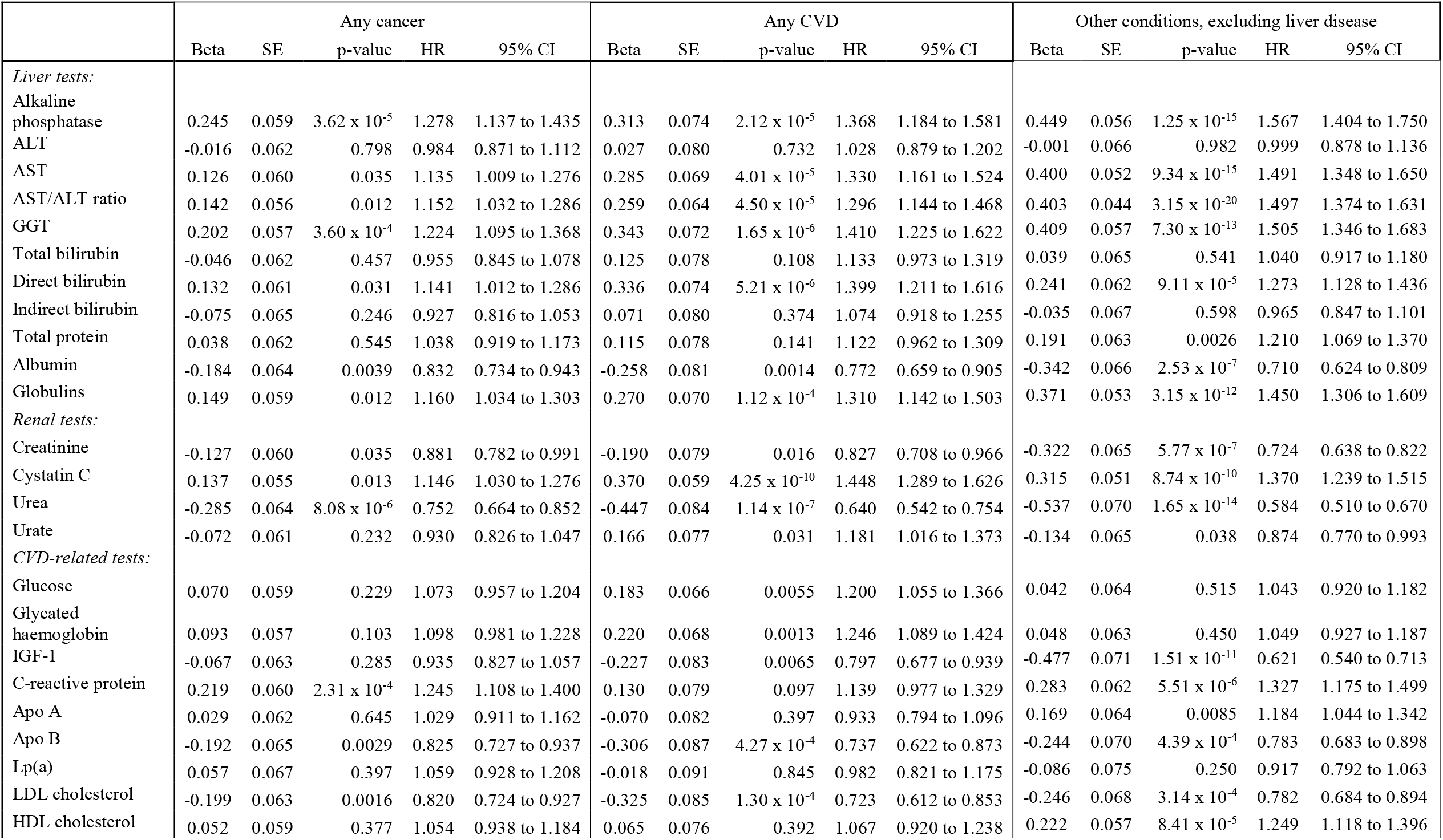
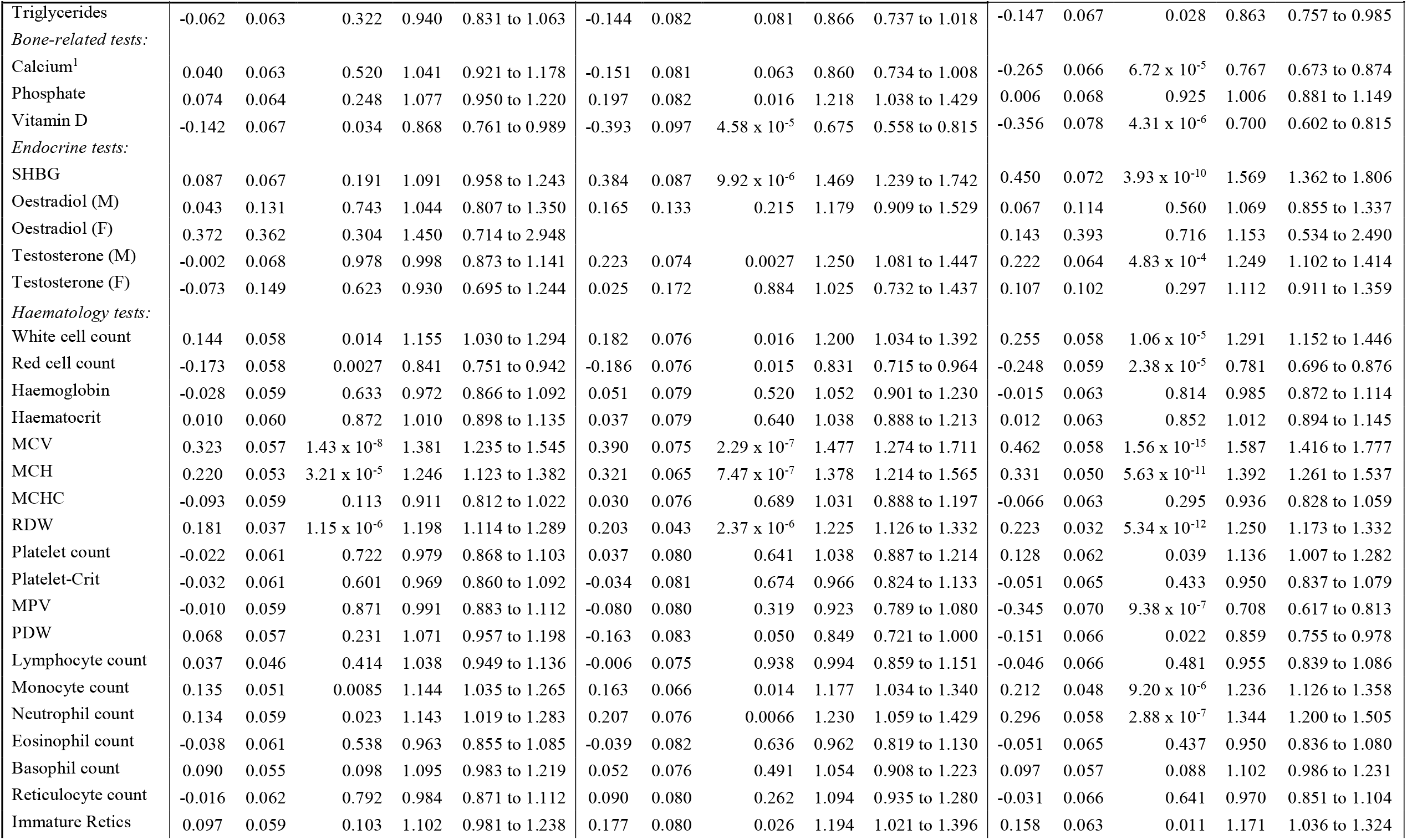
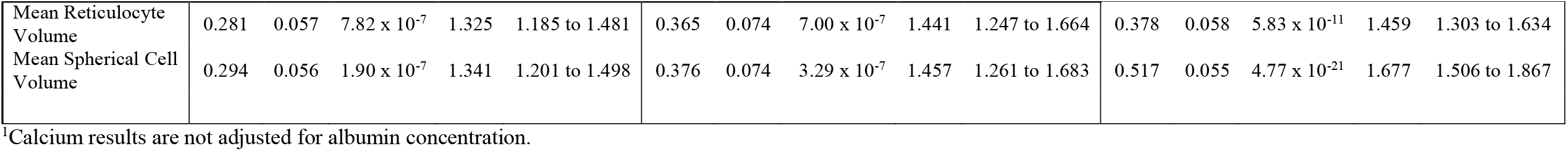
Mortality for major cause-of-death groups in excessive drinkers, by age- and sex-adjusted standardised biomarker results.

**Supplementary Figure 1.**
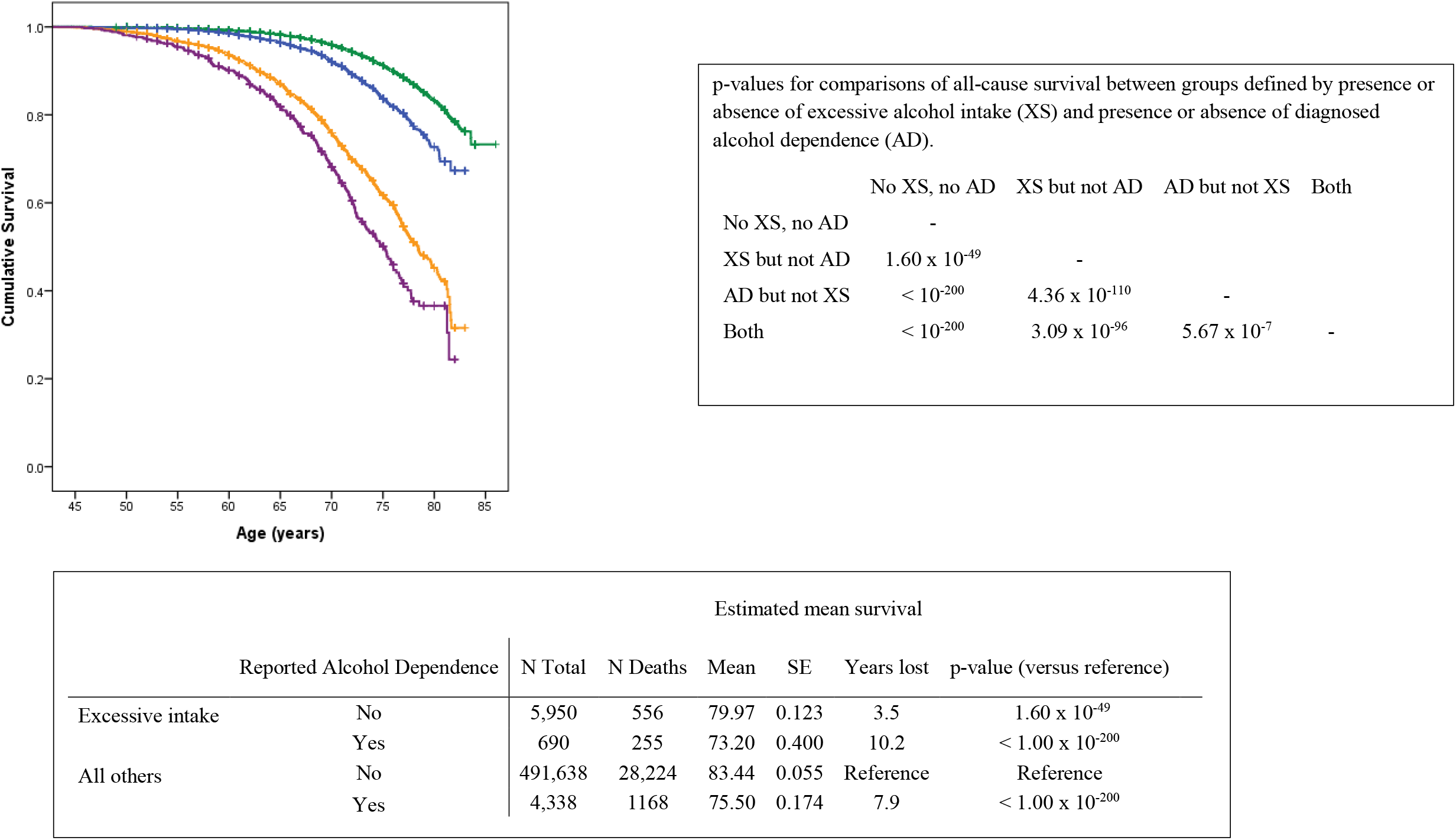
Sex-adjusted Kaplan-Meier mortality curves by excessive drinking and reported diagnosis (from hospital or death records) of alcohol dependence. Green line: no reported excessive drinking, no alcohol-dependence diagnosis; blue line: excessive drinking but no alcohol dependence diagnosis; orange line: no excessive drinking but alcohol dependence diagnosis; purple line: excessive drinking and alcohol dependence diagnosis.

**Supplementary Figure 2.**
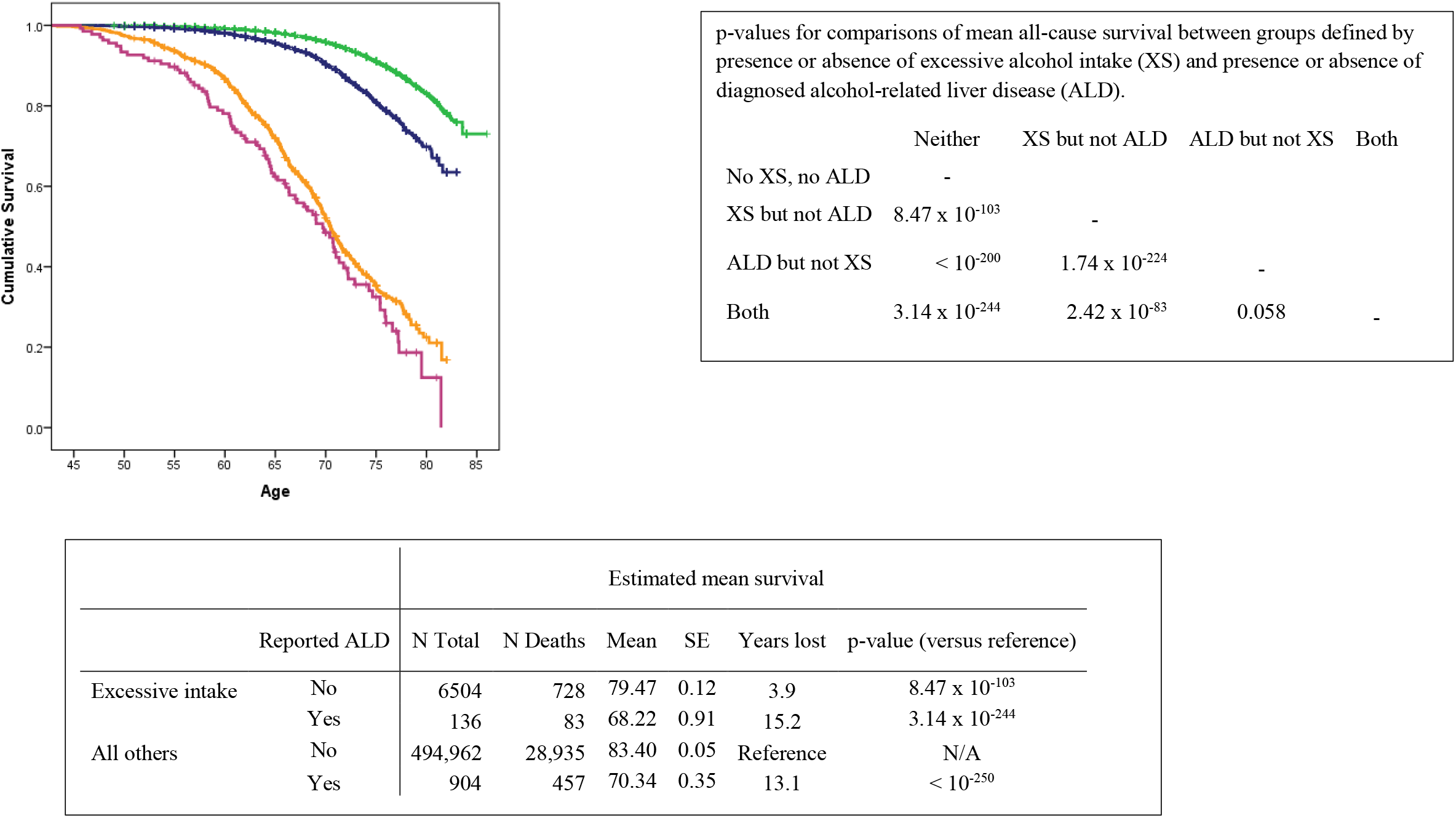
Sex-adjusted Kaplan-Meier mortality curves by excessive drinking and reported diagnosis (from hospital or death records) of alcohol-related liver disease (ALD). Green line: no reported excessive drinking, no ALD diagnosis; blue line: excessive drinking but no ALD; orange line: no excessive drinking but ALD diagnosis; purple line: excessive drinking and ALD.

**Supplementary Figure 3.**
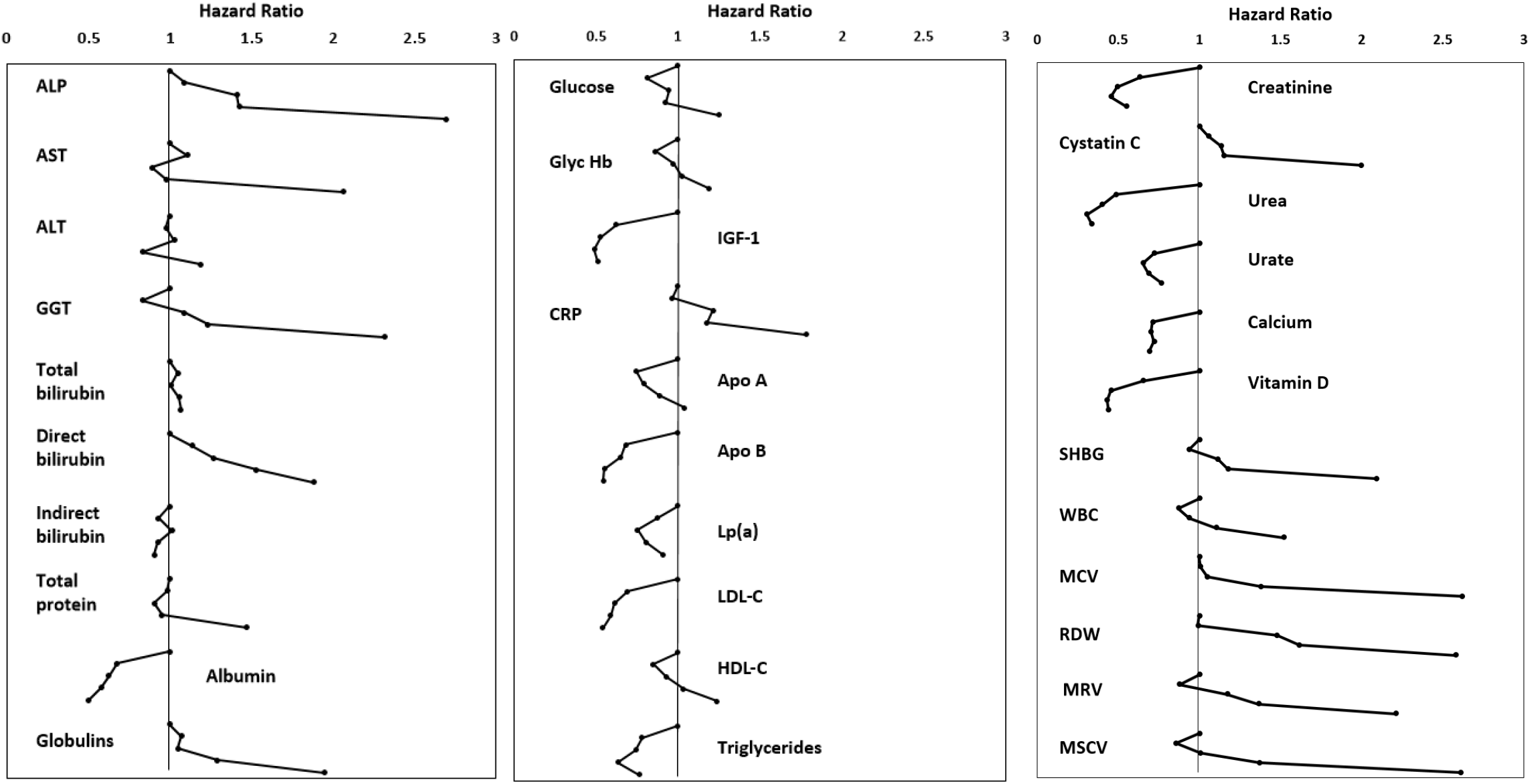
Hazard Ratios (HR) for all-cause mortality by quintiles of biochemical test results, excessive drinkers only. The three panels show results for liver function tests, for tests related to cardiometabolic risk, and for other tests. For each test, the points plotted (moving down the charts) show HRs for quintiles 2, 3, 4 and 5 relative to quintile 1 (the lowest quintile).

## Notes

**Conflict of interest statement** TRM has conducted clinical research with AbbVie, Genfit, Gilead, and Merck but none of these are related to this manuscript.

**Financial support statement** This work was supported, in part, by the National Institutes of Health, USA (U01 AA018389). Funders had no role in study design, data analysis, conclusions, writing and revision of the paper or decisions on publication.

### Competing Interest Statement

TRM has conducted clinical research with AbbVie, Genfit, Gilead, and Merck but none of these are related to this manuscript.

### Funding Statement

This work was supported, in part, by the National Institutes of Health, USA (U01 AA018389). Funders had no role in study design, data analysis, conclusions, writing and revision of the paper or decisions on publication.

### Author Declarations

Information used in this analysis was obtained from the UK Biobank (application number 18870). Participants gave informed consent, and our application was approved by the UK Biobank through their procedures, consistent with the UK Biobank Ethics and Governance Framework (https://www.ukbiobank.ac.uk/learn-more-about-uk-biobank/about-us/ethics, accessed 2022-03-22).

